# Rare coding mutations identify 36 large-effect risk genes in obsessive-compulsive disorder and chronic tic disorders

**DOI:** 10.1101/2025.10.10.25337672

**Authors:** Belinda Wang, Matthew N. Tran, Sheng Wang, Yuting Liu, Emily Olfson, George Wang, Nawei Sun, Jeanselle Dea, Charles Ochieng’ Olwal, Lyvia Bertolace, Michael H. Bloch, Carolina Cappi, Yi-Chieh Chang, Denise Chavira, Barbara J. Coffey, Martha J. Falkenstein, Adam C. Frank, Martin E. Franklin, Stephanie Garayalde, Helena Garrido, Marco Grados, Rami Hatem, Allyna-London Howell, Starlette Khim, Jennie M. Kuckertz, Mindy M. Le, Allison Libby, Ryan J. McCarty, Mary E. McNamara, Daniel McNeil, Euripedes C. Miguel, Cara Nasello, Binh Nguyen, Tenzin Norbu, Lauren Oh, Ashley Ordway, Catherine Paciotti, Viviana A. Peskin, Christopher Pittenger, Helen Blair Simpson, Heather Simpson Martin, Max A. Tischfield, Jinchuan Xing, Jessica J. Zakrzewski, Tourette International Collaborative Genetics (TIC Genetics), Andrea Dietrich, Donald L. Gilbert, Pieter J. Hoekstra, Young Shin Kim, Samuel Kuperman, Alyssa Rosen, Samuel H. Zinner, Mehdi Bouhaddou, Robert A. King, Guy Rouleau, Kerry J. Ressler, Carol A. Mathews, Nevan J. Krogan, Nenad Sestan, Jay A. Tischfield, A. Moses Lee, Gary A. Heiman, Thomas V. Fernandez, A. Jeremy Willsey, Matthew W. State

## Abstract

Obsessive-compulsive disorder (OCD) and chronic tic disorders (CTD) are highly heritable. Recent progress in OCD genomics has highlighted small-effect common alleles. Rare mutations have previously been found to carry large risks for OCD and CTD but only four high-confidence (hc) genes have been identified. We analyzed whole-exome sequencing data from 3,964 individuals with OCD, CTD, or both, including 2,418 trios. We find an excess in cases of *de novo* and rare protein-damaging mutations and identify 36 hc genes (false discovery rate [FDR] < 0.1), four of which overlap with OCD GWAS loci. Risk genes are shared among OCD, CTD, autism spectrum disorder, and other neurodevelopmental conditions. Transcriptomic and network analyses highlight mechanistic convergence and increased risk gene expression in postnatal cerebellum and pre- and postnatal cortex and striatum. Dozens of large-effect OCDCTD genes offer insights into pathogenesis and a path forward for illuminating pathophysiology and identifying novel treatment targets.

## Introduction

Obsessive-compulsive disorder (OCD) and chronic tic disorders (CTD), including Tourette syndrome (TS) are neuropsychiatric conditions that affect 1-2% of the population^1,2^, conferring substantial morbidity and societal burdens^3–7^. Both OCD and CTD are highly heritable^8,9^. However, findings from genetic studies to date have not substantially clarified pathogenesis, disease mechanisms, or led to progress in the clinic, including in treatment development.

Clinically, OCD is characterized by intrusive thoughts or images (obsessions) and repetitive behaviors or mental acts (compulsions) performed to alleviate distress, typically beginning in late childhood or early adulthood^4,5^. CTD is defined by persistent, recurrent motor movements and/or vocalizations, often preceded by premonitory urges^6,7^, with typical onset in early childhood, peak severity around 10-12 years, and improvement during adolescence^10^. Severe cases can result in life-long impairment, and co-occurring neuropsychiatric conditions including attention-deficit/hyperactivity disorder (ADHD), OCD, and mood disorders are common^6,7,11^.

OCD and CTD frequently co-occur. 50% of individuals with CTD exhibit obsessive-compulsive symptoms, and 20–30% of those with OCD have a history of tics^11,12^. The two disorders share substantial heritability^13^ and neuroimaging, clinical, and animal studies implicate dysfunction of cortico-striatal-thalamo-cortical (CSTC) circuitry, which governs action selection and inhibitory control, in both conditions^14,15^. These parallels suggest overlapping etiologies, though differing natural histories and treatment responses also point to the potential for disorder-specific pathogenic and pathophysiological mechanisms.

Given the marked heritability of OCD and CTD, efforts to define their genetic bases have been longstanding^9,16^. Similar to other prevalent medical and psychiatric conditions, common alleles of small effect explain a substantial proportion of the population risk for both conditions^17,18^. Recent genome-wide association studies (GWAS) identified 30 OCD loci from 53,660 cases^19^, but only a single CTD locus from 9,619 cases^20^, reflecting the modest effect sizes of common risk alleles in both conditions and the need for larger cohorts to power future studies. The identification of rare, pathogenic coding variants provides a complementary strategy to leverage genomics to gain insight into these disorders^21^, and such variants have been confirmed to contribute to both OCD and CTD^22–26^. To date, however, only four genes identified via whole-exome sequencing (WES) have reached well-accepted significance thresholds (FDR < 0.1) and are considered hc: two each for OCD (*CHD8, SCUBE1*)^24^ and CTD (*CELSR3, WWC1*)^23^.

For several impairing neurodevelopmental disorders (NDD), including autism spectrum disorder (ASD), progress in identifying heterozygous mutations of large effect has enabled studies of construct-valid animal models, revealed spatio-temporal and mechanistic convergence across risk genes^21^, and is now serving as the basis for clinical trials of mechanism-driven treatments^27^. Motivated by the potential for similar progress in OCD and CTD, we analyzed new and previously reported WES data from 3,964 probands affected with OCD, CTD, or both conditions—2,418 from parent-child trios and 1,546 singletons, roughly doubling prior case sample sizes^23,25^. We compared these findings to 1,734 control trios. Analysis of the disorders separately yields 12 hcOCD and 10 hcCTD genes. Combining OCD and CTD probands yields 34 OCDCTD hc genes. Two genes identified as hc in the disorder-specific analyses meet the threshold for a probable (p) risk gene (FDR ≥ 0.1 and < 0.3) when the cohorts are combined.

All told, our analyses yield 36 hc genes associated with one or both disorders. Comparisons with hc genes from other NDDs, including ASD^28–32^, developmental delay and/or intellectual disability (DD/ID)^33^, and schizophrenia (SCZ)^34^, reveal both shared and distinct risks. This pattern is recapitulated in transcriptomic analyses, which show elevated risk gene expression during prenatal cortical development, similar to ASD, but also distinctively in later postnatal stages, consistent with the relatively later onset of OCD and CTD. These analyses also find statistically significant increased expression in telencephalic projecting excitatory neurons. Finally, analysis of gene interaction databases shows OCD and CTD genes exhibit significantly increased connectivity. These findings, along with the results of both bulk brain and single-cell expression studies, point to spatiotemporally convergent biological processes and pathways underlying the etiology and pathogenesis of OCD and CTD.

## Results

### Cohort overview and study design

We analyzed new and previously sequenced WES data from individuals with OCD, CTD, or both, and neurotypical controls (Fig. 1a; Supplementary Note 1; Supplementary Tables 1–3). After quality control, the case cohort included 2,418 parent–proband trios and 1,546 singletons. The control cohort included 1,734 trios of neurotypical siblings and their parents from the Simons Simplex Collection^35^. Parents of European ancestry were used as singleton controls (n = 2,380). In total, 872 trios (36%) and 1,080 singletons (70%) were newly sequenced for this study.

**Figure 1.**
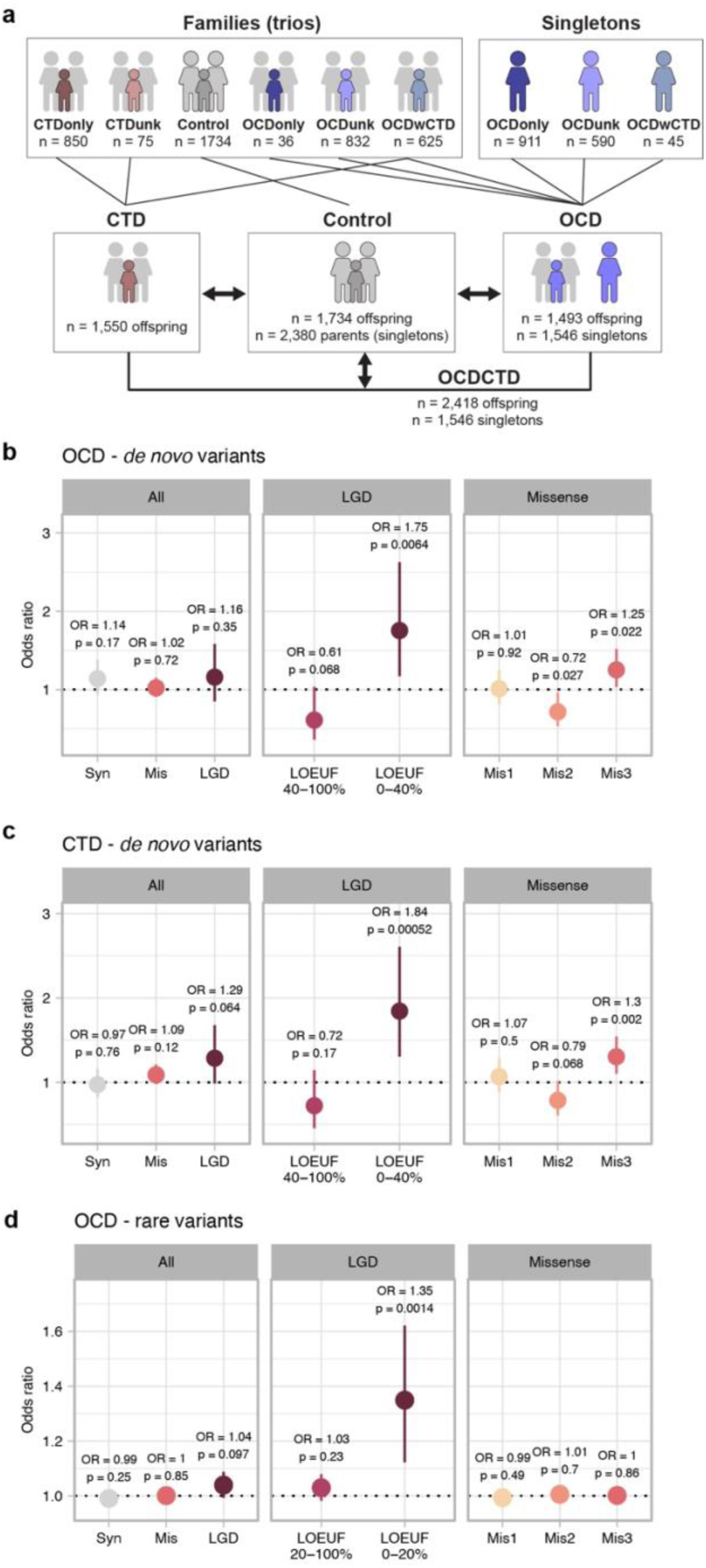
Gene discovery overview and *de novo* and rare variant burden analysis. **a,** Cohort overview. Shown are the numbers of families (affected offspring with two unaffected parents) and singletons (affected individuals without available parental data) contributing to trio-based and singleton analyses. Samples were classified by diagnostic status as CTDonly (CTD without OCD), CTDunk (CTD with unknown OCD status), OCDonly (OCD without CTD), OCDunk (OCD with unknown CTD status), or OCDwCTD (co-occurring OCD and CTD), and grouped into OCD, CTD, or combined OCDCTD cohorts for gene discovery analyses. **b–d**, Case versus control enrichment for *de novo* (dn) variants in OCD trios (b), CTD trios (c), and rare variants in OCD singletons (d). Odds ratios (OR) and 95% confidence intervals from logistic regression are shown. Consistent with established convention^22–24,30,34,37^, *p*-values are not corrected for multiple tests. Allele frequency thresholds in a population reference^36^ were ≤ 0.05% for dn variants and ≤ 0.005% for singleton rare variants (see Methods). Results are shown for (left panels) “All” variants (synonymous, missense, and likely gene-disruptive [LGD]), (middle panels) “LGD” variants stratified by genic constraint (lower LOEUF deciles indicate higher constraint), and (right panels) “Missense” variants classified by predicted deleteriousness (Mis3 = most deleterious^38^).

Trio probands represented a range of diagnostic subtypes, including OCD without CTD (OCDonly, n = 36), OCD with co-occurring CTD (OCDwCTD, n = 625), OCD with unknown CTD status (OCDunk, n = 832), CTD without OCD (CTDonly, n = 850), and CTD with unknown OCD status (CTDunk, n = 75) (Fig. 1a; Supplementary Table 2). All case singletons carried an OCD diagnosis (OCDonly, n = 911; OCDunk, n = 590; OCDwCTD, n = 45; Supplementary Table 3).

We processed all WES, both newly generated and previously published, through a unified pipeline with standardized alignment and variant-calling (Methods; Supplementary Fig. 1). Quality control confirmed reported family structures and excluded samples that failed sequencing quality metrics or showed unexpected relatedness (Supplementary Figs. 2 and 3; Supplementary Tables 4 and 5). All singleton analyses were restricted to individuals of European ancestry to minimize confounding from population structure. To reduce artifacts from sequencing coverage differences, analyses were limited to coding regions meeting specified quality metrics across all sources (Methods).

Building on prior work in OCD and CTD^22–25^ and studies of other NDDs^29,30,33,34^, we evaluated three classes of rare variation (see Methods): (1) rare *de novo* mutations in families (“dn”; allele frequency [AF] ≤ 0.05% in a population reference database^36^), (2) rare transmitted variants in families (“transmitted”, AF ≤ 0.005%) and (3) rare variants in singletons (“rare”; AF ≤ 0.005%). Given the absence of parental data, dn versus transmitted variants could not be differentiated among singletons. In trio probands, we identified 4,028 dn variants (Supplementary Table 6) and 308,078 transmitted variants, and in singletons, we identified 296,736 rare variants.

Prior sequencing studies of NDDs and neuropsychiatric conditions, including OCD and CTD, showed that risk-contributing dn and rare variants are damaging to protein function and enriched among genes constrained against disruptive mutations^22–25,29,30,33,34,37^. Therefore, we categorized variants by predicted functional impact: (1) likely gene-disrupting (LGD, including frameshift, stop-gain, and essential splice site changes) and (2) missense variants predicted to be the most damaging (missense 3 or Mis3)^38^.

LGD variants were further stratified by gene-level “<u>L</u>oss-of-function <u>O</u>bserved/<u>E</u>xpected <u>U</u>pper bound <u>F</u>raction (LOEUF)” scores^39^, where lower scores indicate stronger intolerance to loss of function (LoF) mutations. While prior work^22–25^, including related to ASD^29,30^, served as a useful reference for establishing functional impact parameters, we set specific threshold criteria for constraint in the current study based on our empirical data (see Supplementary Note 2). We refer to the combined set of LGD and Mis3 variants as damaging (Dam) variants. Synonymous variants (Syn) were used as a control.

### Rare damaging variants contribute to OCD and CTD risk

To confirm whether various classes of rare variation, including dn, transmitted, and/or rare variants, contribute to OCD and CTD, we compared variant frequencies between cases and controls. We applied two complementary analytic approaches: logistic regression to assess whether individuals carrying at least one variant were more likely to be cases (Fig. 1b-d; Supplementary Fig. 4d), and Poisson or negative binomial regression to evaluate whether cases carried more variants overall (Supplementary Fig. 4a-c,e; Methods). Reported *p*-values are unadjusted, following the convention established in prior WES studies^22–24,30,34,37^.

Across all cohorts (OCD, CTD, and OCDCTD), cases showed elevated rates of dnLGD in LoF-intolerant genes, specifically among the lowest four LOEUF deciles. We similarly found elevated rates of dnMis3 variants in cases versus controls, consistent with prior WES studies^22–25^ (Fig. 1b and c; Supplementary Fig. 4). We observed no differences in transmitted LGD or transmitted missense variants in trios, even within the lowest LOEUF decile (Methods; data not shown). OCD singletons showed a higher frequency of rare LGD variants in the two lowest LOEUF deciles, in line with previous work^25^ (Fig. 1d; Supplementary Fig. 4). Synonymous variants did not differ between cases and controls (Fig. 1b–d; Supplementary Fig. 4). Based on these results, we included dnLGD, dnMis3, and rare LGD variants in subsequent gene discovery analyses (Supplementary Note 2, Supplementary Figs. 5–7, and Supplementary Tables 7, 8, and 10).

Various lines of genetic evidence have long suggested shared risk between OCD and CTD^24,40^. Consequently, we assessed the overlap of genes carrying dnDam variants across three trio cohorts of similar size: OCDwCTD, CTDonly, and OCDonly/unk (OCDonly and OCDunk combined due to the small size of OCDonly). Permutation testing revealed significantly greater overlap than expected by chance between CTDonly and OCDonly/unk, and between OCDonly/unk and OCDwCTD (Fig 2a; Supplementary Table 9). No significant overlap was observed with controls.

**Figure 2.**
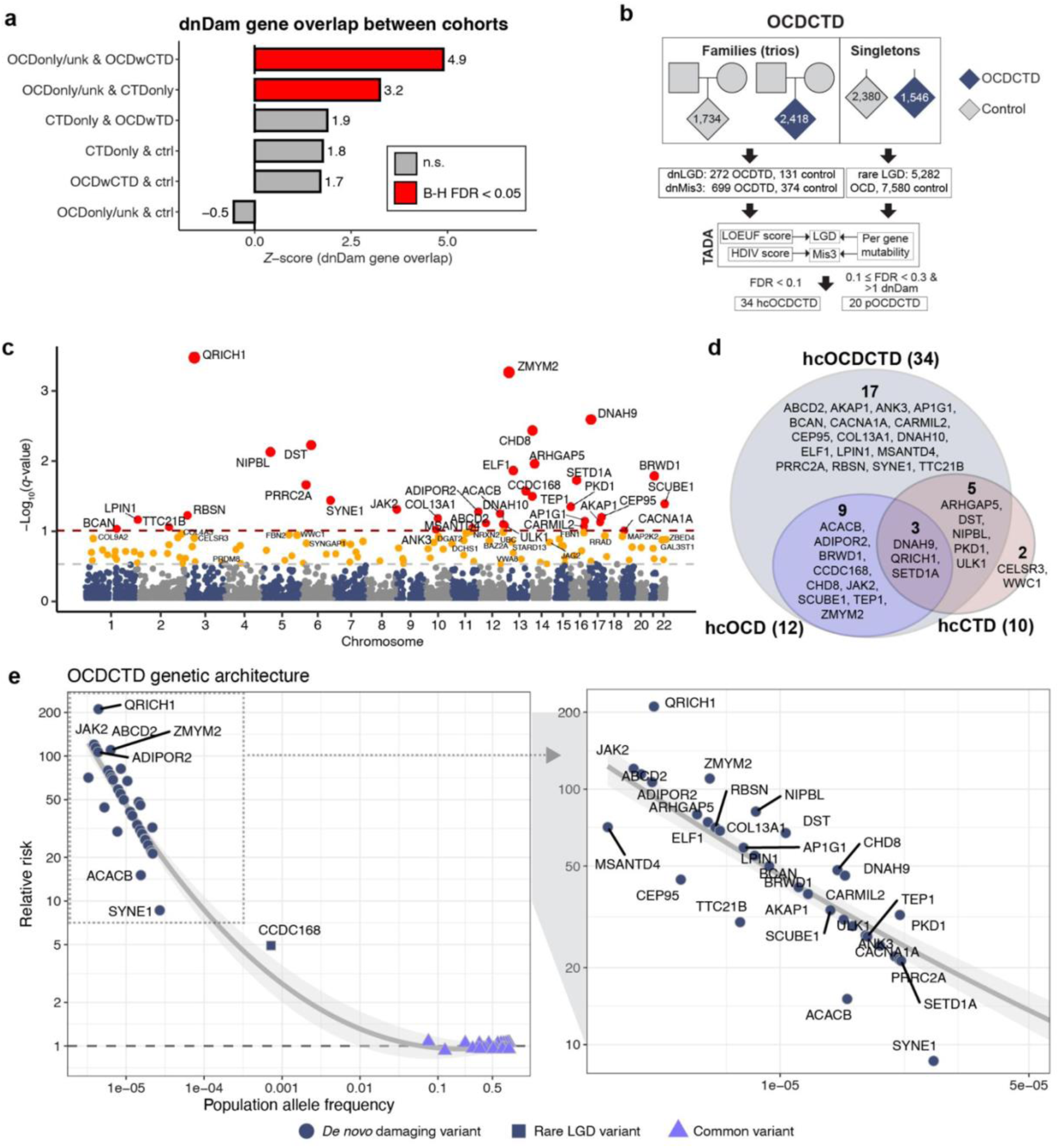
Genetic association results for OCD, CTD, and OCDCTD. **a,** Overlap of genes with dnDam variants between cohort pairs to assess shared genetic risk between OCD and CTD. Cohorts include offspring with CTD without OCD (CTDonly), co-occurring OCD and CTD (OCDwCTD), OCD without CTD or with unknown CTD status (OCDonly/unk), and neurotypical controls (ctrl). One-sided *z*-scores indicate the extent to which the observed gene overlap exceeds chance, based on 10,000 random permutations. Bars are colored by Benjamini– Hochberg (BH)-corrected FDR < 0.05 (red), not significant (n.s., gray). **b**, Sequencing data for the OCDCTD cohort was analyzed using a Bayesian framework (TADA^42^) that incorporated predicted deleteriousness of likely gene-disrupting (LGD) and missense variants, and gene-specific mutability. Total variants observed per cohort are shown. Genes with FDR < 0.1 were defined as high-confidence (hc); those with 0.1 ≤ FDR < 0.3 and >1 dnDam variants were defined as probable (p). **c**, Manhattan plot of –log₁₀(*q*) by chromosomal position for the OCDCTD cohort. Dashed lines denote FDR thresholds of 0.1 and 0.3. Points are colored by FDR (red: < 0.1, orange: ≥ 0.1 and < 0.3), with labels shown for hc and p genes only. **d**, Overlap among hcOCD, hcCTD, and hcOCDCTD risk genes. Numbers indicate gene counts. **e**, Population allele frequency versus relative risk (RR) for hcOCDCTD risk genes and OCD GWAS loci^13^. Shapes indicate the variant class used to estimate relative risk (RR). RR reflects the case-to-control frequency ratio or GWAS odds ratio. LOESS curve with 95% CI shown.

CTD has previously been shown to have a strong male:female sex bias^6^. Male predominance in OCD has been identified only in childhood-onset cases^4,41^. Among our trio probands, the observed male:female ratio was 3.8:1 for CTD and 1.4:1 for OCD. A prior study reported elevated dnLGD rates in LoF-intolerant genes among males with OCD^25^. Using Poisson regression, we observed no sex-specific differences in dn variant rates in any variant class, though power was limited by sample size (Methods; Supplementary Fig. 8; Supplementary Tables 11 and 12).

Using Maximum Likelihood Estimation (MLE), we estimated that 550 OCD, 1,094 CTD, and 760 OCDCTD genes contribute to risk through dn and rare variants. These results are robust to an alternative analytic method (Methods, Supplementary Table 13). Based on the observed excess frequency in cases versus controls, we estimate that 19–50% of damaging dn and rare variants likely contribute to OCD and/or CTD (Supplementary Table 14). At the individual level, we estimate 7–8% of OCD and/or CTD probands carry at least one risk-contributing dnDam variant, and 3.0% of OCD singletons carry a rare LGD variant (Supplementary Table 15).

### Identification of large-effect risk genes

To identify genes associated with OCD and/or CTD, we applied a well-established Bayesian framework, <u>T</u>ransmission <u>A</u>nd *<u>D</u>e novo* <u>A</u>ssociation (TADA)^30,42^, to integrate evidence from dnLGD, dnMis3, and rare LGD variants (Methods; Fig. 2b; Supplementary Fig. 9). TADA uses variant class– specific priors informed by the estimated effect sizes for each class to improve power for gene discovery. For each autosomal protein-coding gene, we calculated a Bayes factor (BF) summarizing the cumulative evidence for association, accounting for gene-specific mutation rates and the estimated number of contributing risk genes. Consistent with prior studies^22–24,29,43^, genes with FDR < 0.1 were designated hc, and those with FDR ≥ 0.1 and < 0.3 were classified as p. However, given the identification of several p genes based on only a single dnDam mutation, we elected to restrict the p designation in the current study to those genes meeting the p FDR threshold while also carrying at least 2 dnDam variants (Supplementary Tables 16 and 17).

Using these definitions, the OCD cohort alone identifies 12 hc- and 16 pOCD genes, while the CTD cohort alone yields 10 hc- and 6 pCTD genes. An omnibus analysis of both cohorts together identifies 34 hc- and 20 pOCDCTD genes (Fig. 2c,d; Supplementary Fig. 9; Supplementary Tables 16 and 17). In total, 36 genes meet hc criteria in at least one analysis (OCD, CTD, or OCDCTD). Two hcCTD genes, *CELSR3* and *WWC1*, identified in the disorder-specific analysis no longer meet the hc threshold in the combined OCDCTD analysis. Their reclassification as p coincides with combining OCD and CTD cohorts that included both trios and singletons. For both genes, we observe an increased rate of rare LGD mutations in singleton OCD controls versus cases. However, both genes remain hc in an analysis of OCDCTD restricted to trios. Similarly, four hcOCDCTD genes (*ABCD2*, *DNAH9*, *DNAH10*, and *TEP1*) also show higher rates of rare LGD mutations in singleton controls versus cases. Further investigation of these genes confirms that three (*ABCD2, DNAH9,* and *DNAH10*) are hc regardless of whether the OCDCTD analysis includes only trios or both trios and singletons. *TEP1* is a p gene in the trio-only OCDCTD analysis and hc in the combined OCDCTD analysis (Supplementary Table 16, Supplementary Note 3).

Given that our study included many previously reported samples, we did not seek to perform an independent replication of published results. However, our current findings do not alter the designation of the hcOCD genes *CHD8* and *SCUBE1*^24^ or the hcCTD genes *CELSR3* and *WWC1*^22,23^. We also identified a new dnMis3 variant in *WWC1* from an OCDwCTD proband not included in our previous CTD-focused studies^22,23^. In contrast, several previously reported genes are not identified as hc or p in our study (see Supplementary Note 4). This includes *SLITRK5*, which previously approached genome-wide significance^25^ in an OCD case-control analysis. Of note, the reported prior association was supported by transmitted rare Mis3 variants in families and rare Mis3 variants in singletons. Neither of these variant classes showed significantly elevated rates in cases versus controls in our dataset and, therefore, were not used for gene discovery in the current study (Fig. 1d; Supplementary Fig. 4c).

Collectively, hc genes identified in our study carry very large effects, with odds ratios (ORs) indicating up to 210-fold increases in risk (57-fold on average), contrasting with the very modest effects typically found for common risk alleles, including in OCD^13^ (ORs less than 1.1; Fig. 2e; Supplementary Fig. 10; Supplementary Table 18). In the current study, the gene *CCDC168* is identified solely through rare LGDs and confers intermediate risk (OR = 4.9). This finding is consistent with the general expectation that rare transmitted NDD-associated LGD variants will carry effect sizes falling between those associated with dnDam variants and those of common alleles.

To explore how the various phenotypes contributed to our gene discovery, we decomposed Bayes factors (BFs) to determine which cohort subgroups (OCDonly, OCDunk, OCDwCTD, CTDonly, or CTDunk) contributed to the identification of each of the 36 hc genes (Methods). 30/36 (83.3%) genes have evidence for association derived from both OCD and CTD cases (Fig. 3a; Supplementary Fig. 11; Supplementary Table 19). Only one gene (*CESLR3*) is supported exclusively by mutations in CTDonly individuals. Five genes (*BRWD1, CACNA1A, CCDC168, CHD8,* and *JAK2*) are supported only by variants in OCD cases without CTD or with unknown CTD status (OCDonly and OCDunk). We also assessed the relative contributions of various variant classes and find that dnLGD and dnMis3 mutations from trios contribute more evidence for gene discovery than rare LGD variants detected in singleton cases (Fig. 3b; Supplementary Fig. 11; Supplementary Table 19). Several genes are supported solely by dnLGD mutations (*ARHGAP5*, *BRWD1, ELF1*, *PRRC2A,* and *SETD1A*); one gene is supported only by dn missense mutations (*NIPBL*), and as noted, one gene is supported only by rare LGD variants in singletons (*CCDC168*).

**Figure 3.**
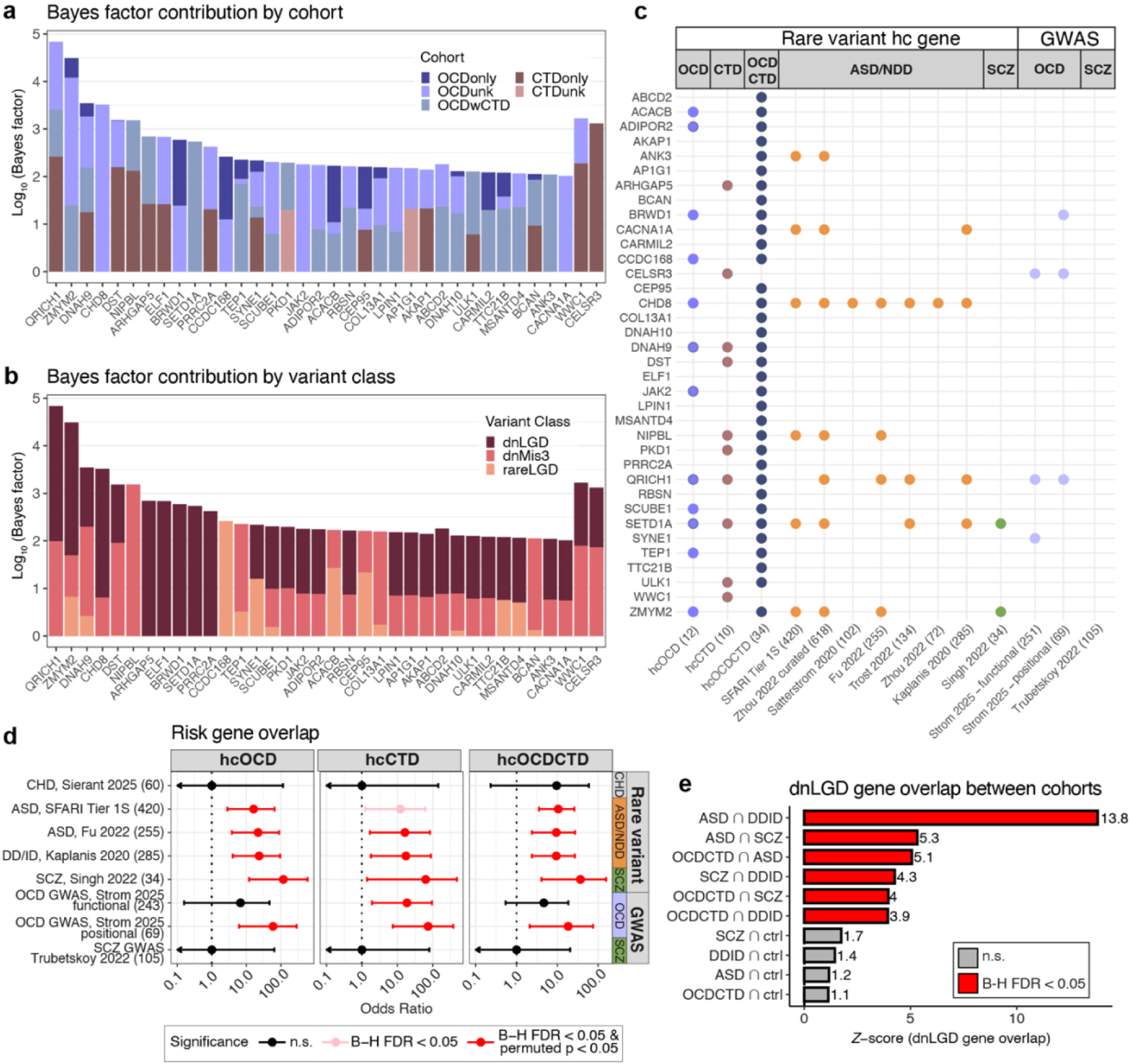
Risk gene profiles and overlap with other neurodevelopmental disorders. **a,b,** Contribution of phenotype (a) and variant class (b) to the statistical evidence (quantified by Bayes factor^42^) for each of the 36 high-confidence (hc, FDR < 0.1) risk genes identified in OCD, CTD, or OCDCTD. **c,** Overlap of the hc genes identified in this study with previously published rare variant–associated gene sets for autism spectrum disorder and neurodevelopmental disorders (ASD/NDD) and schizophrenia (SCZ), and genes that map to OCD and SCZ GWAS loci^13,28–34,82^ (see Methods). Colored dots indicate gene set membership, publications are given by first author and year, and gene set sizes are in parentheses. **d,** Enrichment of hcOCD, hcCTD, and hcOCDCTD genes in previously reported neuropsychiatric gene sets. Odds ratios (ORs) and 95% confidence intervals from two-sided Fisher’s exact tests comparing hcOCD, hcCTD, and hcOCDCTD genes to external gene sets, including ASD/NDD, SCZ, congenital heart defects (CHD), and OCD GWAS loci^13,28–34,45^. Gene set sizes are indicated in parentheses. Colors: not significant (n.s., black), Benjamini–Hochberg [B-H] FDR < 0.05 (pink), B-H FDR < 0.05 and empirical p < 0.05 (10,000 permutations; red). ORs for sets with zero overlap were set to 1 for visualization. **e**, Overlap of genes with dnLGD variants between OCDCTD, ASD, DD/ID, SCZ, and neurotypical control (ctrl) cohorts^30,33,34^. One-sided *z*-scores indicate the extent to which observed gene overlap exceeds chance, based on 10,000 random permutations. Bar colors: B-H FDR < 0.05 (red), n.s. (gray).

### Shared genetic risk for OCD and CTD derives from both common and rare variants

The largest OCD GWAS meta-analysis to date identified 30 genome-wide significant loci^13^. To increase our ability to detect overlap between rare and common variants, we also considered 31 additional loci reported as significant in subgroup GWAS analyses of cohorts ascertained by different methods^13^, for a total of 61 loci. We compiled two risk gene sets–one based on functional or regulatory connections to the 30 genome-wide significant loci (“functional”), and one based on physical proximity to the 61 loci (“physical”)–and compared these to the 36 hc genes identified in the present study (see Methods). Four hc genes (*BRWD1, CELSR3*, *QRICH1,* and *SYNE1*) are present in both the rare variant and common allele gene sets, exceeding chance expectations (Fig. 3c,d; Supplementary Table 20). In line with this, gene-level <u>M</u>ulti-marker <u>A</u>nalysis of <u>G</u>eno<u>M</u>ic <u>A</u>nnotation (MAGMA)^44^, which tests whether aggregated single-nucleotide polymorphism (SNP) associations implicate a gene, shows nominal enrichment for seven hc genes, including *CELSR3* and *QRICH1* (Supplementary Fig. 12; Supplementary Note 5). As noted, in our study, *CELSR3* dn mutations were observed exclusively in CTDonly individuals, and the gene is an hcCTD gene in disorder-specific analyses, an hcOCDCTD gene in a trio-only analysis, and a pOCDCTD gene in the omnibus analysis of singleton and trios. The overlap observed with genome-wide significant OCD GWAS results^13^ broadly supports the association with the OCDCTD phenotype and suggests that further evaluation of *CELSR3* expression isoforms and the functional impact of transmitted variants may help clarify our rare variant findings.

### Shared genetic risk with neurodevelopmental disorders

Based on results from studies of rare variant risk in ASD, SCZ, and DD/ID, we suspected that the rare variant risks identified in the present study might lead to a wide range of NDDs^28–34^. We therefore tested two complementary hypotheses: 1) that dn mutations found among affected individuals with OCD and/or CTD would also be present in ASD, SCZ and/or DD/ID cohorts, and 2) that hcOCDCTD risk genes would overlap with established ASD, DD/ID and SCZ risk genes.

First, we find that genes carrying dnLGD mutations in the current study significantly overlap with those identified in studies of ASD, DD/ID, and/or SCZ (*z*-scores 3.9–5.1 by permutation testing, Supplementary Fig. 3e; Supplementary Fig. 17; Supplementary Table 21). We next examined hc genes from the current study and similarly observe significant overlap with ASD, DD/ID, and SCZ risk genes identified in prior WES studies and curated databases^28–34^ (Fig. 3c,d; Supplementary Table 20). No overlap is observed with congenital heart disease (CHD) genes^45^ (*p* = 1).

We next sought to more finely characterize the nature of the overlapping risk for our newly identified OCDCTD gene set. To do so, we evaluated gene-level FDR values for all the identified hc genes with respect to their association with OCDCTD and compared these to results from recent large-scale WES studies of ASD^30^, DD/ID^30,33^, and SCZ^34^ (Methods; Supplementary Note 5; Supplementary Fig. 14; Supplementary Table 22). Although the early stage of rare variant gene discovery and incomplete phenotypic data in studies of NDDs preclude definitive conclusions, these analyses suggest that it may well be possible in the future to identify a subset of OCDCTD-predominant or OCDCTD-specific genetic risks.

### Spatial, temporal, and cellular mapping of risk genes

We evaluated the spatial, temporal, and cellular expression patterns of OCD and CTD risk genes using multiple transcriptomic atlases^46–52^. Because the resolution of transcriptomic data varied across resources, we relied on different OCD and/or CTD gene sets for these analyses. To query high-resolution microarray and bulk RNA-seq datasets, we relied on smaller and more stringently defined sets of risk genes, namely hc and p genes. Alternatively, to increase the power of exploratory analyses, we relied on a larger set of genes carrying at least one dnLGD mutation to query sparser single-cell RNA-seq datasets.

Using GTEx bulk RNA-seq data from 54 human tissues^46^, we ranked genes by tissue specificity^53^ and performed ranked gene set enrichment analysis (Methods). Expression in the cerebellum is significantly increased after accounting for multiple testing correction. In addition, nominal enrichment is observed in cortex (frontal and anterior cingulate), striatum (caudate, putamen, nucleus accumbens), amygdala, and peripheral nervous system (tibial nerve) (Fig. 4a; Supplementary Table 23). No enrichment is detected in the 40 non-nervous system tissues. The findings in cerebellum have not previously been reported based on OCD and CTD GWAS, whereas the nominal enrichment noted above aligns with prior analyses^13,20^.

**Figure 4.**
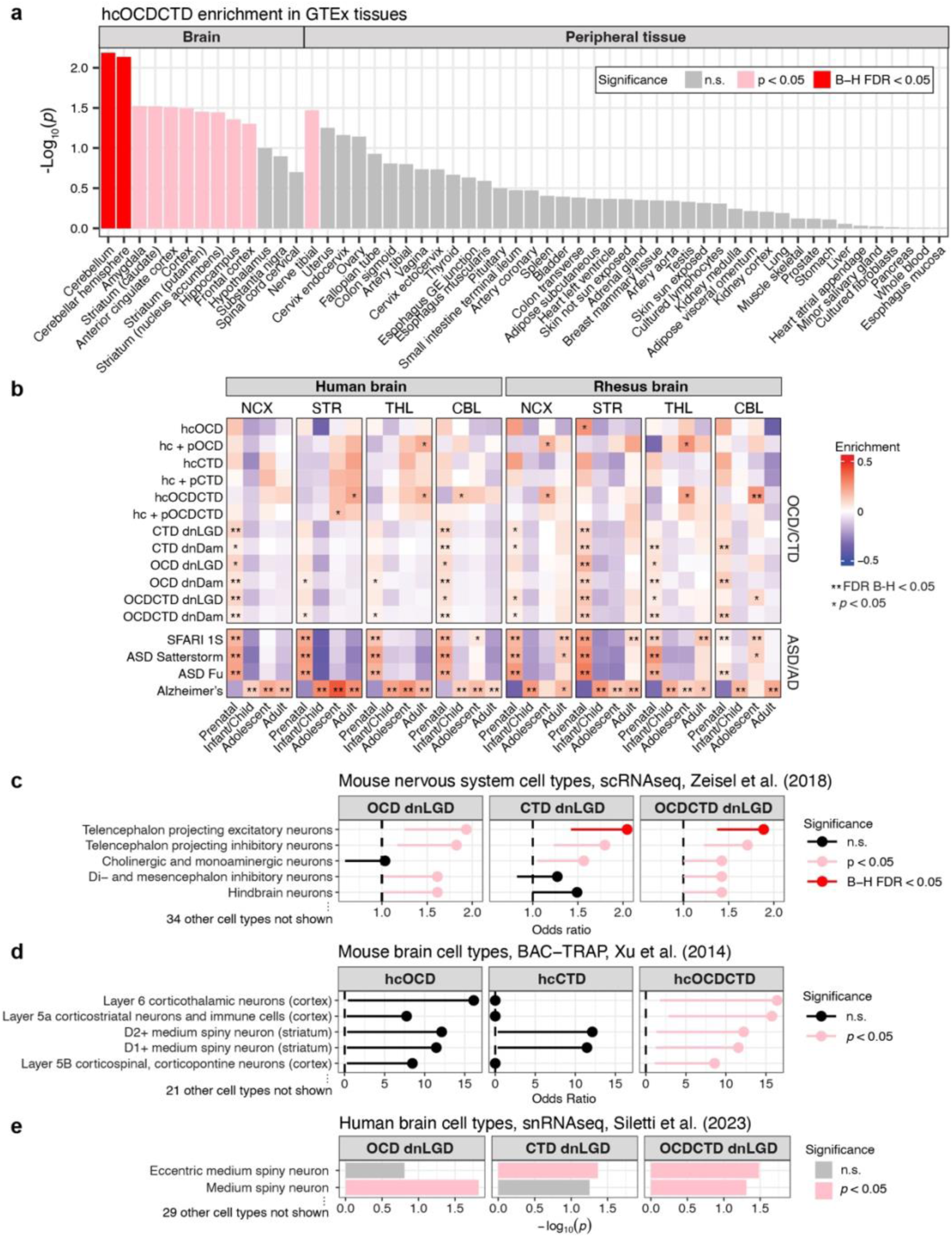
Regional, developmental, and cell type risk gene enrichment. **a,** Enrichment of hcOCDCTD genes across 54 GTEx human tissues^46^. **b,** Developmental expression of hc, the union of hc and p (hc + p), dnLGD (genes with ≥1 observed dnLGD), and dnDam (genes with ≥1 observed dnDam) gene sets in human^48^ and rhesus macaque^49^ neocortex (NCX), striatum (STR), thalamus (THL), and cerebellum (CBL). Heatmaps show enrichment relative to all other genes (one-sided Wilcoxon rank-sum test). HcASD^29,30^ gene sets were included as a control for prenatal-biased expression, and an Alzheimer’s disease^54^ (AD) gene set as a control for postnatal-biased expression. **B-H FDR < 0.05; *nominal *p* < 0.05. **c-e,** Risk gene set enrichment among the top 10% cell type– specific genes from 39 mouse postnatal nervous system cell types^50^ (c), mouse postnatal brain cell type–specific markers defined by BAC–TRAP^51,83^ (d), and 31 adult human brain cell types^52^ (e). To balance specificity and power, smaller hc gene sets were queried in d (higher-resolution microarray data), and larger dnLGD sets in c and e (sparser single-cell data). Only cell types with significant enrichment in at least one gene set are shown. Color coding (a,c–e): red, B-H FDR < 0.05; pink, nominal p < 0.05; gray, not significant (n.s.). Enrichment was calculated by gene set enrichment analyses using expression specificity ranks (a,e) or one-sided Fisher’s exact tests with odds ratios and 95% confidence intervals shown (c,d).

We next evaluated spatial and temporal expression of OCD and CTD risk genes using the BrainSpan human brain atlas, which includes both microarray^48^ and RNA-seq^47^ data, alongside a rhesus macaque atlas^49^ as a primate comparator. Our analyses focused on the neocortex, striatum, thalamus, and cerebellum across prenatal to adult stages; regions were selected based on the identification of adequate sample sizes, with the neocortex being best powered. For each region, we computed gene-level temporal specificity *z*-scores and used one-sided Wilcoxon rank-sum tests to assess whether risk genes showed greater stage-specific expression than background genes (Methods). ASD^28–30^ and Alzheimer’s disease (AD)^54^ were used as controls, given established expression patterns for known risk genes and alleles. Consistent with the results from GTEx, hcOCDCTD genes show elevated postnatal expression in the cerebellum (Fig. 4b; Supplementary Fig. 15). Across regions, OCD-, CTD-, and OCDCTD-associated dnLGD and dnDam genes show prenatal enrichment, with several regions and time points remaining significant after multiple-testing correction (FDR < 0.05). Hc and p risk genes show nominally elevated postnatal expression, particularly in the human striatum and thalamus, a pattern distinct from ASD (Fig. 4b; Supplementary Fig. 15; Supplementary Table 24).

To identify relevant cell types, we mapped risk genes to cell type-specific markers from postnatal transcriptomic atlases. Using a mouse nervous system single-cell RNA-seq atlas^50^, we tested whether dnLGD genes were overrepresented among the 10% most cell type-specific genes^55^ using one-sided Fisher’s exact tests. Across 39 cell types, telencephalic projecting excitatory neurons, which include various subclasses (layer 2–6 cortical pyramidal neurons, hippocampal CA1/subicular neurons, and basolateral amygdalar neurons), are significantly enriched after multiple testing correction (Fig. 4c; Supplementary Fig. 16; Supplementary Table 25). Nominal enrichment is also observed in telencephalic projecting inhibitory neurons (mainly dorsal and ventral striatal D1+ and D2+ medium spiny neurons), hindbrain, di-/mesencephalic inhibitory, and cholinergic/monoaminergic neurons. We further corroborated these findings with hc genes and an orthogonal mouse bacterial artificial chromosome–translating ribosome affinity purification (BAC-TRAP) dataset, which profiled ribosome-bound transcripts across 26 genetically defined nervous system cell populations via microarray^51^ (Fig. 4d; Supplementary Fig. 16; Supplementary Table 26). Our results are consistent with prior OCD and CTD GWAS-based analyses^13,20,50^, which found that common variants were most enriched in telencephalic projecting excitatory and inhibitory neurons for OCD, and in telencephalic projecting excitatory and di-/mesencephalic inhibitory neurons for CTD.

Finally, we evaluated a human adult brain single-nucleus RNA-seq atlas that profiled over 3 million nuclei and identified 31 cell types^52^. Genes were ranked by cell type specificity, defined as the proportion of each gene’s expression attributable to a given cell type. We applied ranked gene set enrichment analysis to assess whether genes carrying dnLGD mutations were overrepresented among the most cell type–specific genes and find nominal enrichment in two medium spiny neuron subtypes (Fig. 4e, Supplementary Fig. 16; Supplementary Table 27).

### Functional connectivity of OCD and CTD risk genes

We evaluated the molecular and functional properties of OCD and CTD risk genes using complementary approaches. Gene Ontology (GO) analysis^56^ of combined hcOCDCTD and pOCDCTD genes reveals overrepresentation in neurodevelopmental processes, with axonogenesis (GO:0007409) remaining significant after multiple-hypothesis correction (*q* = 0.0145; Supplementary Fig. 17; Supplementary Table 28).

We assessed risk gene network connectivity using PCNet^57^, which integrates multiple types of functional interactions, including protein interactions, co-expression, and regulatory relationships. For each hc and p gene set, we measured (1) within-set connectivity, defined as the proportion of risk genes that directly interact with at least one other risk gene in the set, and (2) interactome overlap, defined as the proportion of risk gene pairs sharing more interactors than expected by chance, out of all possible risk gene pairs (Methods; Fig. 5a). The results were compared to null distributions from random gene sets to calculate *z*-scores, where *z* > 1.64 corresponds to one-sided *p* < 0.05. We find significantly elevated within-set connectivity for hcOCD (8/12, *p* = 0.0038), hcCTD (7/10, *p* = 0.021), and hcOCDCTD (24/34, *p* = 0.0001; Fig. 5b; Supplementary Fig. 18a,b; Supplementary Table 29). The interactome overlap results follow a similar pattern, with significant overlap for all three gene sets (all *p* < 0.001; Fig. 5c; Supplementary Fig. 18c; Supplementary Table 30).

**Figure 5.**
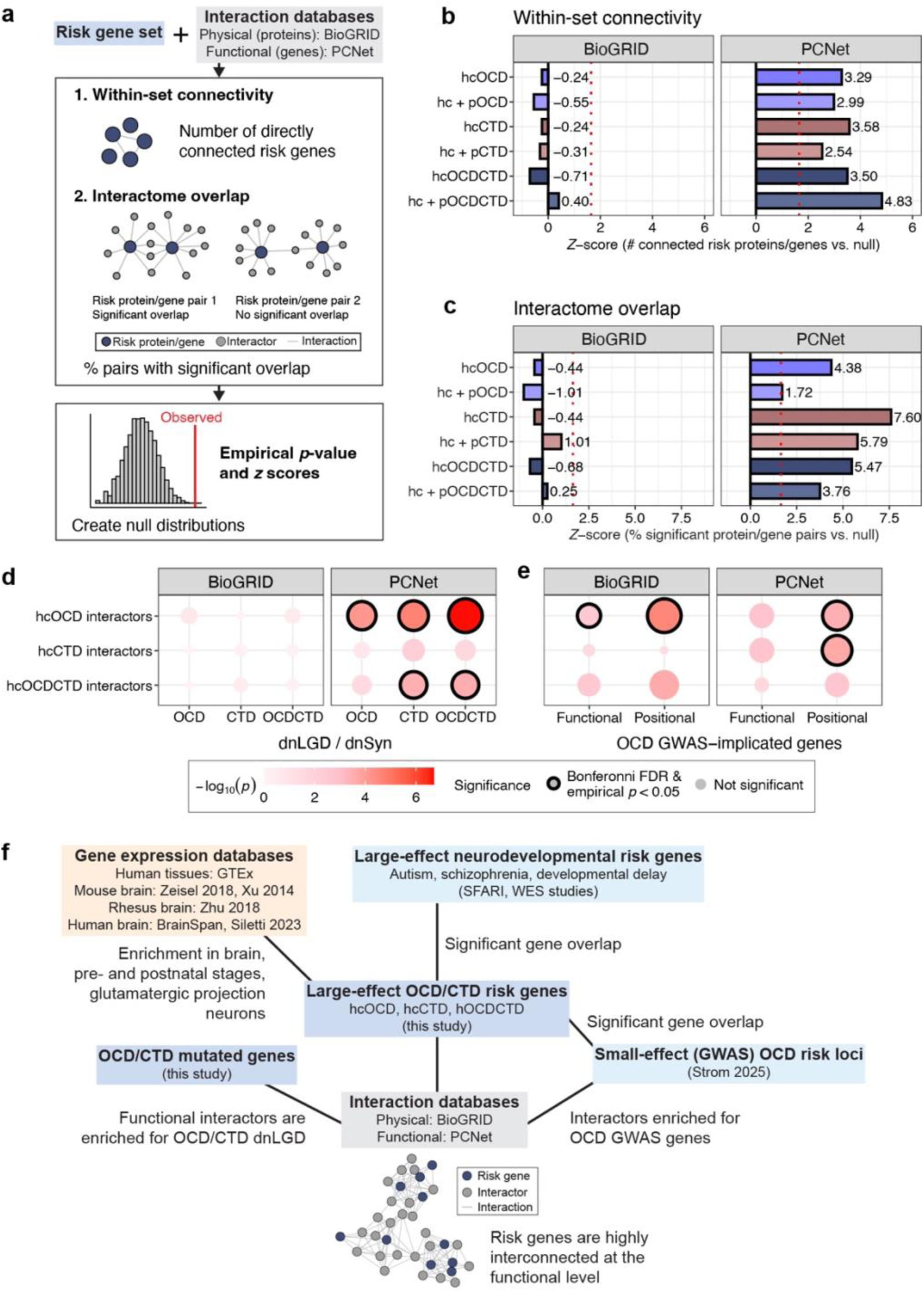
Network analysis reveals functional convergence of OCD and CTD genetic risk. **a,** Framework for assessing connectivity among risk proteins/genes in two interaction networks: BioGRID (physical protein-protein interactions, PPIs), and PCNet (multimodal functional gene interactions, including PPIs)^57,59^. We quantified (1) within-set connectivity, defined as the number of directly interacting risk proteins/genes, and (2) interactome overlap, defined as the proportion of risk protein/gene pairs sharing significantly more interactors than expected in each network. **b,c,** *Z*-scores for within-set connectivity (b) and interactome overlap (c) across networks and various risk protein/gene sets (color coded). Red lines indicate the one-sided threshold for *p* < 0.05 (*z* > 1.64). **d,e,** Enrichment among the first-degree (direct) interactors of hc and hc+p risk proteins/genes in BioGRID and PCNet for (d) dnLGD versus dn synonymous (dnSyn) variants observed in OCD, CTD, or OCDCTD trio probands, and (e) genes mapped to OCD GWAS loci^13^. Dot size and color indicate –log₁₀(*p*) from two-sided Fisher’s exact tests; black outlines denote significance (Bonferroni-adjusted *p* < 0.05 and permutation-derived empirical *p* < 0.05). **f,** Summary of genetic overlap, tissue and cell type specificity, and functional convergence findings.

We next examined whether first-degree interactors were enriched for pathogenic mutations (Methods; Figs. 5d and e; Supplementary Tables 31 and 32). HcOCD and hcOCDCTD interactors are significantly enriched for case dnLGD variants (Fisher’s exact test, Bonferroni-adjusted *p* < 0.01; empirical *p* < 0.01). Interactors of hcOCD and hcCTD also overlap significantly with genes mapped to OCD GWAS loci^13^ (Fisher’s exact test, Bonferroni-adjusted *p* < 0.005; empirical *p* < 0.05). MAGMA gene set-level analysis shows nominal enrichment of OCD common-variant risk among PCNet interactors of hcOCD, hcCTD, and hcOCDCTD (Supplementary Fig. 18d; Supplementary Table 33).

We then explored the functional roles of hc genes using network propagation, which distributes signals from “seed” genes across the PCNet interaction network to quantify connectivity with other genes. Analyses were performed separately with hcOCD, hcCTD, and the union of hcOCD/hcCTD/hcOCDCTD as seed genes. GO overrepresentation analysis with the top 100 genes most significantly connected to the seed sets reveals enrichment of Hippo signaling and “establishment or maintenance of cell polarity” for hcCTD, consistent with our prior CTD WES study^23^, telomere- and DNA biosynthesis-related terms for hcOCD, and “microtubule-based movement” and cilia-related terms across all hc seed sets, consistent with our prior work on ASD risk genes, including the hcOCDCTD gene *CHD8*^58^ (Supplementary Fig. 19, Supplementary Table 34).

We performed similar analyses using BioGRID^59^, which catalogs only physical protein–protein interactions (PPIs). Using this dataset, we do not replicate our findings (Fig. 5a-e; Supplementary Fig. 19; Supplementary Tables 29-33). However, we note that this relatively sparse resource was likely underpowered for the present analyses, and that recent empirical protein interaction studies point to substantial limitations in the currently available public PPI databases, including BioGRID^60^. In contrast, the consistency of findings across multiple different approaches in the PCNet-based analyses suggests that OCD and CTD risk genes do form functionally coherent networks, with their direct interactors enriched for both rare and common variant risk.

## Discussion

The genetic contribution to OCD and CTD has been well established for decades^8,9^, but the identification of specific risk genes and alleles has so far had limited impact on treatment development or the understanding and clinical management of these syndromes. In the case of OCD, successful GWAS studies are navigating the well-known challenges of pursuing many very-small-effect, common non-coding variants that act simultaneously to increase risk^13,61^. Moreover, for CTD, only a single significant GWAS finding has previously been confirmed^20^. Similarly, to date, only a handful of rare copy number variations^62,63^ and large-effect genes^23–25^ have been identified. Against this backdrop, the current work represents a considerable advance. Altogether, the analyses noted above identifies 36 hc genes from separate and combined studies of OCD and CTD cohorts, 34 of which meet hc thresholds in the omnibus OCDCTD analysis.

Our findings indicate that dn and rare pathogenic mutations are carried by approximately 3-8% of affected individuals with OCD and CTD, and these can confer very large effects. Hc risk genes show an average odds ratio of 57 and carry increased risks up to 210-fold. These findings also lay the foundation for a range of tractable experiments aimed at deepening the understanding of these conditions, including studies of construct-valid genetic models^64^ in other species and human induced pluripotent stem cell-based systems such as cerebral organoids^65^, evaluation of protein-protein interactions^60^, and experimental and informatic approaches aimed at identifying mechanistic and spatio-temporal convergence across the growing list of large-effect risk genes^60,66–69^.

Our results confirm substantial genetic overlap between OCD and CTD, evidenced by shared gene-level associations and excess sharing of genes with dn damaging variants across well-phenotyped subgroups, consistent with their clinical similarity and frequent co-occurrence^70,71^. We also observe statistically significant overlap in rare and common variant risk, with 4 hc genes overlapping OCD GWAS loci^72^. While still preliminary, these findings point to the potential for shared pathobiology between rare and common forms of these disorders, which, in turn, raises the prospect of overlapping mechanistically-informed treatment approaches. These findings also underscore the importance of future studies investigating the full spectrum of genetic variation to clarify how rare and common variants act independently and jointly to influence risk.

We also find that OCDCTD risk genes overlap with ASD, DD/ID, and SCZ, but not CHD, in line with extensive evidence that a broad range of neuropsychiatric disorders share genetic risks^61^. Notably, however, some OCDCTD risk genes show no association with ASD or DD/ID, despite the hundreds of large-effect hc genes identified for these NDDs^30,33^. Although further gene discovery will be necessary to power definitive analyses, these results suggest that additional studies could both identify shared origins of clinically overlapping conditions and provide an avenue to elaborate mechanisms and treatments distinctive for OCDCTD. Preliminary transcriptomic analyses support this view, showing similar patterns of expression enrichment in the prenatal brain in OCDCTD and ASD, along with evidence for distinctive postnatal enrichment in OCDCTD that is largely absent in ASD.

Our transcriptional analyses further show that OCD and CTD risk genes are highly expressed in the cortex, striatum, and thalamus during both pre- and postnatal development and the cerebellum during postnatal development. Definitive assessment of region- and stage-specific expression is currently limited by available transcriptomic datasets, which have small sample sizes outside the neocortex.

Future studies will undoubtedly be aided by both expanded developmental expression datasets and additional gene discovery in OCD and CTD. Notably, the most stringently defined analyses presented here, including hc genes, show elevated postnatal brain expression, while the broader set of genes identified by the presence of a dnLGD mutation shows prenatal enrichment patterns similar to those observed with hcASD. These differences likely reflect both biological factors and power limitations, as hc sets are smaller and enriched for higher-penetrance genes. Supporting the idea that risk spans both pre- and postnatal stages, both the hc and broader set of genes showed elevated expression in similar cell types. These include postnatal cortical excitatory projection neurons and striatal medium spiny neurons, consistent with the CSTC circuit implicated in OCD and CTD through neuroimaging, neuromodulation, animal models, and common variant studies^73–79^.

In addition to these circuit-level insights, our network propagation analysis highlights several mechanistic themes–including cell polarity and microtubule and cilia biology–that align with prior CTD findings^23^ and have also emerged prominently in our work in ASD^80,81^, which we show here shares genetic risk with OCDCTD. The consistency of our results with prior mechanistic themes identified in related NDDs, as well as the agreement with existing circuit-level hypotheses, is reassuring and highlights the potential value of the present work, as the discovery of many distinct large-effect genetic etiologies for OCD and CTD offers multiple tractable avenues to deepen our understanding using a range of developmental and systems neuroscience approaches.

As noted, though we find multiple lines of evidence demonstrating increased connectivity and functional convergence among OCDCTD risk genes, a commonly used PPI database, BioGRID^59^, does not replicate these findings. This discrepancy may reflect the fact that PCNet captures functional relationships relevant for OCDCTD that extend beyond direct PPIs. Alternatively, the absence of a signal may simply point to the inherent limitations of existing publicly available PPI resources. For example, our recent study mapping protein interactions encoded by 100 hcASD genes^60^, the largest systematically derived PPI dataset for any NDD to date, found that nearly 90% of identified interactions were absent from existing databases, including BioGRID. These results suggest that studying the interactions of the full set of proteins encoded by the 36 hc risk genes, including in relevant cell types suggested by our expression analyses, along with the assessment of the impact of pathogenic mutations in these interactions, will be valuable in elaborating the OCDCTD proteome and deepening our understanding of mechanistic convergence, disorder pathogenesis, and potential treatment targets.

Overall, the present findings offer novel insights into the genetic architecture of obsessions, compulsions, and tics, confirm and extend key neurobiological hypotheses, and offer promising avenues to leverage large-effect coding mutations, as well as both the overlap and divergence of resulting phenotypes, to elaborate pathophysiology. Perhaps most importantly, our results introduce a ten-fold increase in the number of *bona fide* large-effect risk genes into a rapidly advancing scientific landscape. The pace of progress in neuroscience, functional genomics, proteomics, cell and structural biology, and machine learning is breathtaking, matched by accelerating efforts in mechanism-driven therapeutics research in neuropsychiatric disorders. Together, these developments are positioning the field to deliver on the promise of translating gene discovery into novel and more effective therapeutics for individuals affected by severe neuropsychiatric disorders such as OCD and CTD.

## Consortia

<u>The members of the Tourette International Collaborative Genetics (TIC Genetics) consortium (in</u> <u>alphabetical order):</u> Juliane Ball, Noa Benaroya-Milshtein, Kate Bornais, Keun-Ah Cheon, Barbara J. Coffey, Andrea Dietrich, Erik M. Elster, Dana Feldman, Thomas V. Fernandez, Carolin Fremer, Donald L. Gilbert, Danea Glover, Tammy Hedderly, Gary A. Heiman, Isobel Heyman, Pieter J. Hoekstra, Hyun Ju Hong, Chaim Huijser, Christina Kappler-Friedrichs, Young Key Kim, Young Shin Kim, Robert A. King, Nadine Kirchen, Carolin Sophie Klages, Samuel Kuperman, Bennett L. Leventhal, Holan Liang, Maria Loreta Lopez, Osman Malik, Marieke Messchendorp, Dararat Mingbunjerdsuk, Pablo Mir, Astrid Morer, Kirsten R. Müller-Vahl, Alexander Münchau, Laura Muñoz-Delgado, Tara L. Murphy, Cara Nasello, Kerstin J. Plessen, Veit Roessner, Alyssa Rosen, Guy Rouleau, Simon Schmitt, Chitra Shukla, Sara Sopena, Matthew W. State, Tamar Steinberg, Zsanett Tarnok, Joshua K. Thackray, Meitar Timmor, Jay A. Tischfield, Max A. Tischfield, Anne Uhlmann, Ana Vigil-Pérez, Frank Visscher, Susanne Walitza, Belinda Wang, Sheng Wang, A. Jeremy Willsey, Jinchuan Xing, Samuel H. Zinner.

## Acknowledgements

We are deeply grateful to all the patients and families who participated in the many studies that have contributed data to this manuscript. This work would not have been possible without the brilliance, tenacity, and collaborative spirit of Thomas Lehner, who served for 16 years at NIMH, first as Chief of the Genomics Research Branch and then as Chief Genomics Advisor. We are also grateful to Robert Malenka for his visionary leadership of the FFOR initiative. We thank James T. McCracken for his thoughtful perspectives. We thank the staff of Sampled, the Yale Center for Genome Analysis, and Genome Quebec, as well as Mayra Aldecoa, Diana Bok, Avi Botwinick, Maria Cruz, Kayla Delapenha, Kristin Fleming, Jennifer Garibaldi, Laura Ibanez-Gomez, Jessica Leuchter, Monocle O. Lima, Adam Lombroso, Daniela Martinez, Andrea River Molina, and Vessela Zaharieva for their support of this work.

## Author Contributions

<u>Conceptualization</u>: B.W., S.Wang, E.O., B.J.C., R.A.K., N.Sestan, J.A.T., G.A.H., T.V.F., A.J.W, M.W.S.; Data Curation: B.W., M.N.T., S.Wang, E.O., G.W., B.J.C., A.C.F., J.X., TIC Genetics (N.B.-M., E.M.E., D.F., C.K.-F., M.T., A.V.-P.), A.D., Y.S.K., R.A.K., G.A.H., T.V.F., A.J.W; <u>Formal analysis</u>: B.W., M.N.T., S.Wang; <u>Funding acquisition</u>: B.J.C., D.L.G., S.Kuperman., A.R., S.H.Z., J.A.T., G.A.H., T.V.F., A.J.W, M.W.S.; <u>Investigation</u>: B.W., M.N.T., S.Wang, Y.L., E.O., G.W., N.Sun, C.O.O., L.B., C.C., Y.-C.C., D.C., M.J.F., A.C.F., M.E.F., S.G., H.G., R.H., A.-L.H., S.Khim., J.M.K., M.M.L., A.L., R.J.M., M.E.M., D.McNeil, B.N., T.N., L.O., A.O., C.Paciotti, V.A.P., C.Pittenger, H.B.S., H.S.M., J.J.Z., TIC Genetics (J.B., N.B.-M., K.-A. C., E.M.E., D.F., C.F., T.H., I.H., H.J.H., C.H., C.K.-F., Y.K.K., N.K., C.S.K., H.L., M.L.L., O.M., M.M., D.Mingbunjerdsuk, P.M., A.Morer, K.R.M.-V., A.Münchau, L.M.-D., T.L.M., K.J.P., V.R., S.Schmitt, C.S., S.Sopena, Z.T., M.T., A.V.-P., F.V., S.Walitza), A.D., D.L.G., Y.S.K., S.Kuperman., A.R., S.H.Z., M.B., R.A.K., G.R., K.J.R., C.A.M., J.A.T., A.M.L., G.A.H., T.V.F., A.J.W; <u>Methodology</u>: B.W., M.N.T., S.Wang, B.J.C., J.A.T., G.A.H., T.V.F., A.J.W, M.W.S.; <u>Project administration</u>: B.W., E.O., B.J.C., M.J.F., M.E.F., J.M.K., J.X., TIC Genetics (D.G., I.H., C.K.-F., C.S., Z.T., A.U.), A.D., R.A.K., K.J.R., J.A.T., A.M.L., G.A.H., A.J.W, M.W.S.; <u>Resources</u>: M.H.B., M.E.F., M.G., D.McNeil, E.C.M., C.Pittenger, H.B.S., J.X., TIC Genetics (K.B., P.M., V.R., A.U.), J.A.T., A.J.W, M.W.S.; Software: B.W., M.N.T., S.Wang, Y.L., G.W., C.O.O., A.J.W; <u>Supervision</u>: B.J.C., M.J.F., J.M.K., M.E.M., TIC Genetics (N.B.-M., I.H., P.M., V.R.), Y.S.K., M.B., K.J.R., N.J.K., N.Sestan, J.A.T., A.M.L., G.A.H., A.J.W, M.W.S.; <u>Validation</u>: B.W., M.N.T., S.Wang, Y.L., G.W., J.D., TIC Genetics (J.K.T.); N.Sestan, J.A.T., A.J.W, M.W.S.; <u>Visualization</u>: B.W.; <u>Writing - original draft</u>: B.W. and M.W.S. with input from M.N.T., S.W., Y.L., E.O., J.A.T., G.A.H., A.M.L., T.V.F., and A.J.W; <u>Writing - review and editing</u>: B.W., M.N.T., S.Wang, Y.L., E.O., G.W., N.Sun, J.D., C.O.O., L.B., M.H.B., C.C., Y.-C.C., D.C., B.J.C., M.J.F., A.C.F., M.E.F., S.G., H.G., M.G., R.H., A.-L.H., S.Khim., J.M.K., M.M.L., A.L., R.J.M., M.E.M., D.McNeil, E.C.M., C.N., B.N., T.N., L.O., A.O., C.Paciotti, V.A.P., C.Pittenger, H.B.S., H.S.M., M.A.T., J.X., J.J.Z., TIC Genetics (J.B., N.B.-M., K.B., K.-A. C., E.M.E., D.F., C.F., D.G., T.H., I.H., H.J.H., C.H., C.K.-F., Y.K.K., N.K., C.S.K., B.L.L., H.L., M.L.L., O.M., M.M., D.Mingbunjerdsuk, P.M., A.Morer, K.R.M.-V., A.Münchau, L.M.-D., T.L.M., K.J.P., V.R., S.Schmitt, C.S., S.Sopena, Z.T., J.K.T., M.T., A.U., A.V.-P., F.V., S.Walitza), A.D., D.L.G., P.J.H., Y.S.K., S.Kuperman, A.R., S.H.Z., M.B., R.A.K., G.R., K.J.R., C.A.M., N.J.K., N.Sestan, J.A.T., A.M.L., G.A.H., T.V.F., A.J.W, M.W.S.. All authors approved the final version of the manuscript.

## Ethics declarations

E.O. has received research support or grants from the NIH, The Hartwell Foundation, Tourette Association of America, International OCD Foundation, Misophonia Research Fund, Yale Center for Clinical Investigation, and Yale Child Study Center; is chair of the Early Career Investigator Program Committee and serves on the Board of Directors for the International Society of Psychiatric Genetics (unpaid); and is also a member of the Research Committee for the American Academy of Child & Adolescent Psychiatry (unpaid).

M.H.B. has received grant or research support from Therapix Biosciences, Emalex Biosciences, Neurocrine Biosciences, Janssen Pharmaceuticals, and NIH; serves as Associate Editor of *The Journal of Child Psychology and Psychiatry* and on the editorial boards of the *Journal of Child and Adolescent Psychopharmacology and JAACAP*; and has received royalties from Wolters Kluwer for *Lewis’s Child and Adolescent Psychiatry: A Comprehensive Textbook, Fifth Edition*; and moonlighting pay from the Veterans Administration.

In the past three years, C.Pittenger has consulted for Biohaven Pharmaceuticals, Freedom Biosciences, Transcend Therapeutics, UCB BioPharma, Mind Therapeutics, Ceruvia Biosciences, F-Prime Capital Partners, and Madison Avenue Partners; has received research support from Biohaven Pharmaceuticals, Freedom Biosciences, and Transcend Therapeutics; owns equity in Alco.

Therapeutics, Mind Therapeutics, and Lucid/Care; receives royalties from Oxford University Press and UpToDate; and holds patents on pathogenic antibodies in pediatric OCD and on novel mechanisms of psychedelic drugs. None of these relationships are related to the current paper.

In the last three years, H.B.S. has received royalties from UpToDate, Inc.; served on a Scientific Advisory Board for Otsuka Pharmaceuticals (December 12, 2024); and receives a stipend from the American Medical Association for serving as Associate Editor for *JAMA Psychiatry*.

K.R.M.-V. has received financial or material research support from DFG: GZ MU 1527/3–1 and GZ MU 1527/3–2, and Almirall Hermal GmbH; consultant’s and other honoraria from AlphaSights Ltd., Aurora, Canopy, Cansativa, Canymed, DHMS Direct Health Medical Services Ltd./Wellster Healthtech Group, Emalex, Neuraxpharm, Renafan, Sanity Group, Synendos Therapeutics AG, Takeda, and Tetrapharm; is an Advisory/Scientific Board Member for Branchenverband Cannabiswirtschaft e.V. (BvCW), Sanity Group, Synendos Therapeutics AG, and Therapix Biosciences Ltd.; has received speaker’s fees from Ärztekammer Niedersachsen, Bundesverband der pharmazeutischen Cannabinoidunternehmen (BPC), Cogitando GmbH, diaplan GmbH, FomF GmbH, Grow, Laleto GmbH, Landesamt für Soziales, Jugend und Versorgung Mainz, Noema, streamedup! GmbH, VBG – Unfallversicherung Hamburg, Universitätsklinikum Hamburg-Eppendorf, Universitätsklinikum Münster, and WeCann; has received royalties from Elsevier, Medizinisch Wissenschaftliche Verlagsgesellschaft Berlin, and Kohlhammer; is an associate editor for *Cannabis and Cannabinoid Research*; is an Editorial Board Member of *Medical Cannabis and Cannabinoids* and *MDPI-Reports*; and is a Scientific Board Member for *Zeitschrift für Allgemeinmedizin*.

D.L.G. has received compensation for expert testimony for the U.S. National Vaccine Injury Compensation Program, through the Department of Health and Human Services; received payment for medical expert opinions through TeladocHealth International; served as a paid consultant for PTC Therapeutics, Noema Pharma, and Emalex Biosciences and has received travel support to attend investigator meetings; provided educational lectures for Illumina, Inc. and PTC Therapeutics, Inc.; received salary compensation through Cincinnati Children’s for work as a clinical trial site investigator from Emalex Biosciences, Inc. (clinical trial, Tourette Syndrome), PTC Therapeutics (registry and clinical trial, Amino Acid Decarboxylase Deficiency), Neurocrine Biosciences (clinical trial, cerebral palsy), and Quince Therapeutics (clinical trial, ataxia telangiectasia); and received book/publication royalties from Elsevier and Wolters Kluwer.

K.J.R. has performed scientific consultation for Bioxcel, Bionomics, Acer, and Jazz Pharma; serves on Scientific Advisory Boards for Sage, Boehringer Ingelheim, Senseye, and the Brain Research Foundation; and has received sponsored research support from Alto Neuroscience.

N.J.K. has received research support from Vir Biotechnology, F. Hoffmann-La Roche, and Rezo Therapeutics; has a financially compensated consulting agreement with Maze Therapeutics; serves as President and Board member of Rezo Therapeutics; and holds shares in Tenaya Therapeutics, Maze Therapeutics, Rezo Therapeutics, and GEn1E Lifesciences.

N. Sestan is a co-founder, board member, and shareholder of Bexorg.

T.V.F. has received research support or grants from the NIH, Misophonia Research Fund, and Yale Child Study Center; received an honorarium for participation in the 2025 Pediatric Psychopharmacology

Update Institute by the American Academy of Child & Adolescent Psychiatry; and is paid for expert testimony and consultation by DLA Piper LLC.

A.J.W.’s current affiliation is with Calico Life Sciences LLC, South San Francisco, CA, USA.

M.W.S. serves on the Scientific Advisory Board of MapLight Therapeutics and is a shareholder in Bexorg.

All other authors declare no competing interests.

## Data Availability

All datasets are described in the manuscript or its Supplementary Information. Sample metadata are provided in Supplementary Tables 2 and 3, and dn mutations detected in trios are reported in Supplementary Table 6. Summary statistics from the TADA analysis are provided in Supplementary Table 16. For contributing datasets for which individual-level distribution is permitted, the data have been or are being deposited in public repositories (e.g., the NIMH Data Archive (NDA) and/or the database of Genotypes and Phenotypes (dbGaP)). The accession numbers are: TIC Genetics Phases 1 and 2 (BioProject PRJNA384374) and Phase 3 (NDA 2958), TSAICG (BioProject PRJNA384389), OCGAS (dbGaP phs000903.v1.p1), and SSC (NDAR: DOI 10.15154/1149697).

## Code availability

The software and code used are described throughout the Methods. All software packages used in this study are publicly available. In brief, for sequencing data generation, we used GATK v4.2.0 (with Picard v2.25.0), and VerifyBamID2 version v1.0.6. Sample and variant quality control and analyses were performed using Hail 0.2 (https://hail.is/). Additional analyses and visualization were executed in R 4.2.3 using the following key packages: tidyverse_2.0.0, biomaRt_2.54.1, GenomicRanges_1.50.2, fgsea_1.24.0, ggtern_3.5.0, Seurat_4.3.0.1, and ComplexHeatmap_2.15.4. The R code used to generate TADA association results will be publicly available on GitHub at the time of publication: https://github.com/Psychiatry-State-Lab/OCDCTD

## Methods

### Study participants

All adult participants and parents of children provided written informed consent, along with written or oral assent of their participating child. The Institutional Review Board (IRB) of each participating site approved the study. WES data were aggregated from previous OCD studies^24,25^, CTD studies^22,23^, and new samples from TIC Genetics^85^ and a study supported by the Foundation for OCD Research (FFOR). Control samples were drawn from the Simon’s Simplex Collection (SSC)^86–88^, including neurotypical siblings of autism probands and their parents. Neurotypical SSC siblings and parents were used as controls for trio and singleton analyses, respectively. The pre-quality control (QC) dataset comprised 15,716 WES samples (10,261 cases and 5,455 controls). Ascertainment criteria for each cohort are detailed in Supplementary Note 1. Pre- and post-QC sample counts by source are summarized in Supplementary Table 1. Supplementary Tables 2 and 3 provide post-QC sample-level information, including OCD and CTD diagnostic status.

### Sample processing and variant identification

Supplementary Fig. 1 provides an overview of sample and variant QC.

#### Sequencing and alignment

Newly generated CTD and OCD exomes were captured using the Twist Bioscience Comprehensive Exome Panel (Twist Bio) and sequenced on the Illumina NovaSeq 6000 platform (S4 flowcells, 100 bp paired-end reads), targeting a mean coverage of 100x. Exome capture kits and sample metadata for all new and previously published data are provided in Supplementary Tables 2–3. All sequencing data, including previously generated data, were processed using the Genome Analysis Toolkit (GATK v4.2.0) with Picard v2.25.0, following GATK Best Practices^89^. Reads were aligned to the Genome Reference Consortium Human Reference Genome Build 38 (GRCh38) with the Burroughs Williams Aligner (BWA)-MEM algorithm via a SamToFastq > MergedBamAlignment workflow. Duplicate reads were marked with MarkDuplicatesSpark, base quality was recalibrated with BaseQualityScoreRecalibration, and sample contamination was estimated using VerifyBamID2 v1.0.6 (FREEMIX). To account for underestimation, FREEMIX values were divided by 0.75 (https://github.com/gatk-workflows). Adjusted estimates were used as input to GATK HaplotypeCaller to generate per-sample gVCFs.

#### Joint genotyping, hard filters, and variant annotation

Joint genotyping followed GATK Best Practices. We used GenomicsDB to combine all samples, performed joint genotyping with GnarlyGenotyper, and applied Variant Quality Score Recalibration (VQSR), retaining variants with PASS at 99.7% truth-set sensitivity for SNVs and indels. The joint-genotyped gVCF was processed with Hail v0.2. We applied hard filters (specifically: read depth [DP] ≥ 10 and genotype quality [GQ] ≥ 20) and removed variants in low-complexity regions^84^. For *de novo* (dn) analyses, we excluded ChrY variants in females and non-pseudoautosomal region (PAR) ChrX variants in males. Variants were annotated with Ensembl VEP v113^90^ using the GENCODE v47 transcript model. Plugins incorporated PolyPhen-2 HDIV predictions^38^ and allele frequencies from gnomADv4.1 exomes. Additional annotations included gnomAD v2.1.1-based missense badness, Polyphen-2, and constraint (MPC) scores^91^ and gene-level constraint metrics (specifically: LOEUF and pLI) from gnomAD v4.1^36,39,91^. We retained coding variants with the following Variant Effect Predictor (VEP)^90^ consequences: frameshift_variant, stop_gained, splice_donor_variant, splice_acceptor_variant, inframe_deletion, inframe_insertion, missense_variant, start_lost, stop_lost, or synonymous_variant.

#### Defining consensus and callable regions

To minimize biases from differences in the multiple capture platforms, we defined “consensus callable” regions. For trios, we intersected GRCh38 coding regions with targets from seven capture platforms (±100 bp padding) and retained bases with DP ≥ 15 in all family members. For singletons, we intersected GRCh38 coding regions with targets from four capture platforms covering > 95% of samples, retaining sites with DP ≥ 10 and GQ ≥ 20 in ≥ 90% of samples from each of the three most common platforms (each representing at least 10% of cases). These consensus callable regions were used for all burden analyses. For gene discovery, singletons used the same consensus callable region, while trios used a broader “restricted callable” region, defined as the intersection of padded capture targets and bases with DP ≥ 15 in all trio members. This approach ensured uniform coverage while maximizing sensitivity for dn gene discovery, consistent with previous studies^22,23^.

#### Sample-level QC

Sample-level QC was performed in several steps: (1) Relatedness and pedigree checks: using Hail identity_by_descent, we confirmed parent–child relationships (IBD_HAT ≥ 0.35; Z1 ≥ 0.7), identified duplicate samples (IBD_HAT ≥ 0.95), and flagged highly related pairs (0.35 ≤ IBD_HAT < 0.95; Z1 ≤ 0.3). Duplicates were resolved by prioritizing retention of complete trios. (2) Sequencing and exome capture integrity: a genome-wide coverage matrix (∼25k intervals)^92^ was used for principal component analysis (PCA) and clustering. Nine clusters matched known capture kits (Supplementary Fig. 2). Samples that failed to cluster or showed discordant trio clustering—indicative of poor coverage or atypical capture—were excluded. (3) Sex inference and ancestry: sex was inferred with Hail impute_sex; trios without the expected male/female parental pattern were removed. Ancestry was inferred by PCA and a random forest classifier trained on 1000 Genomes superpopulations (sklearn::RandomForestClassifier). Individuals of European (EUR) ancestry (probability ≥ 0.8) were retained for singleton analyses (Supplementary Fig. 3; Supplementary Table 7). (4) Data-specific QC: For trios, we removed outliers with excess rare dn variants (above the 99.7th percentile of a Poisson distribution), leaving 4,136 high-quality trios (2,402 OCD/CTD; 1,734 controls, Supplementary Table 4). In gene discovery analyses, we included dn variants from 16 trios that had been previously Sanger validated but came from trios excluded by our current sample QC approach, for a final cohort of 4,152 trios (2,418 cases, 1,734 controls). For singletons, we analyzed VQSR-PASS variants with ≥ 90% coverage across three main exome kits. Samples were excluded as outliers based on call rate (3 standard deviations [SD] below the mean), singleton count (3SD above the mean), and heterozygous/homozygous ratio or transition/transversion ratio (3SD above or below the mean). After filtering, 3,926 high-quality European singletons remained (1,546 cases, 2,380 controls; Supplementary Table 5).

#### *De novo* variant identification and QC

Dn variants were identified as heterozygous in the proband and as homozygous reference in both parents, with cohort allele frequency (AF) ≤ 0.001, depth (DP) ≥ 15 in all family members, and proband allele balance (AB) 0.3 ≤ AB ≤ 0.7. Candidate variants were scored with DeNovoCNN^93^ and further filtered to AF ≤ 0.0005 across all gnomAD v4.1 exome populations (global, African/African American, Admixed American, Ashkenazi Jewish, East Asian, Finnish, Middle Eastern, Non-Finnish European, South Asian, and unassigned groups). Prior studies have shown that manual *in silico* visualization of sequencing reads is highly concordant with Sanger validation^22–24,94^.

Accordingly, all dn variants not previously Sanger validated were manually reviewed using the Integrative Genomics Viewer (IGV, https://igv.org). Variants were confirmed if present in the proband and absent in both parents. Confirmation rates were 165/176 for indels (93.75%) and 3,065/3,075 for SNVs (99.65%), comparable to recent reported Sanger validation rates (87.5–95.7% for indels; 98.8– 99.5% for SNVs^25,94^). As noted, previously Sanger-validated dn variants^22–25^ from samples that failed current QC were retained for gene discovery analyses but were excluded from burden testing (n = 20 variants across 16 probands, <0.8% of total case dn variants; Supplementary Table 6). Two dn variants that previously failed Sanger validation were excluded from all analyses. Only one dn variant per gene per family was retained, prioritizing the most damaging allele. All dn variants used in burden or gene-discovery (TADA) analyses are listed in Supplementary Table 6.

#### Transmitted rare variant identification and QC

Variants were required to meet general filters (DP ≥ 10, GQ ≥ 20, 0.3 ≤ AB ≤ 0.7), pass variant quality score recalibration (VQSR), and be rare (internal AF across 4,136 trios ≤ 0.0005; global gnomAD AF ≤ 0.00005). These transmitted rare variants were used to calibrate cohort-specific variant quality score log-odds (VQSLOD) thresholds for transmitted and singleton rare variants. Because capture and sequencing platforms differed between cases and controls, SNV VQSLOD thresholds were calibrated separately to avoid systematic bias. We focused on control trios captured with the Roche NimbleGen SeqCap EZ Exome Library, v2 (EZV2); (all controls) and case trios captured with TwistBio (the predominant case platform). Within each group, we enumerated transmitted and untransmitted alleles using Hail’s transmission_disequilibrium_test, focusing on singleton synonymous SNVs (AC = 1 in parents). For each VQSLOD percentile bin, we performed a binomial test under the null hypothesis of 50% transmission and selected the bin with the highest p-value (most balanced transmission)^29,95^. The final SNV VQSLOD thresholds were: VQSLOD_OCD/CTD_ ≥ −1.444 and VQSLOD_SSC_ ≥ −2.177; an indel threshold of VQSLOD ≥ −1.812 (99.5% tranche) was also applied. Variants below these thresholds were removed. Individual-level entries were exported for each trio, with transmitted variants defined as those in which the proband genotype (GT) was 0/1, and untransmitted variants defined as those in which the proband GT was 0/0 and at least one parent had GT 0/1. For burden analyses, additional filtering was applied, requiring DP ≥ 15 and rarity in gnomAD v4.1 exome populations with ancestry-stratified thresholds (AF ≤ 0.00005 in non-Finnish Europeans and AF ≤ 0.0005 in African/African American, Admixed American, Ashkenazi Jewish, East Asian, Finnish, Middle Eastern, South Asian, and unassigned cohorts). Only one rare transmitted variant per gene per offspring was retained, prioritizing the most damaging allele.

#### Singleton rare variant identification and QC

For EUR samples passing QC, we retained heterozygous genotypes (GT = 0/1) with GQ ≥ 20, DP ≥ 10, and AB 0.25–0.75. Variants were required to pass the same VQSLOD thresholds as transmitted rare variants. Rare variants were defined as those with MAC ≤ 5 in the combined case–control cohort and global and non-Finnish European AF ≤ 0.00005 in gnomAD v4.1 exomes. For each gene per individual, only the most damaging variant was retained.

### Burden analyses

#### Variant class definitions and prioritization

We analyzed three classes of genetic variation: dn rare variants (trios), transmitted rare variants (trios), and rare variants with an unknown mode of inheritance (singletons). Within each class, we examined three functional categories: synonymous, missense, and likely gene-disrupting (LGD; stop-gained, frameshift, splice donor/acceptor variants). To enrich for deleterious variants, we stratified LGD and missense variants by gene-level constraint and predicted functional impact, respectively. LGD variants were stratified by LOEUF (Loss-of-function Observed/Expected Upper bound Fraction from gnomADv4.1 constraint metrics) and, in a comparative analysis, pLI (Probability of Loss-of-function Intolerance) scores^95^. Missense variants were stratified using PolyPhen-2 human divergence (HDIV)^38^ into Mis3 (probably damaging), Mis2 (possibly damaging), and Mis1 (benign). We also evaluated an alternative missense classifier using the MPC score^96^, with MPC ≥ 2 defining “MisB” variants. We empirically evaluated the performance of variant classifiers in enriching for deleterious variants in cases versus controls using burden analyses, with synonymous variants serving as a negative control to confirm model calibration (Supplementary Figs. 5–7; Supplementary Tables 7–10). Full stratification criteria and the rationale for selecting variant classes to utilize in gene discovery are provided in Supplementary Note 2.

#### De novo, transmitted, and rare variant burden (exome-wide)

To evaluate the contribution of rare coding variants to disease risk, we assessed variant burden between cases and controls (Fig. 1; Supplementary Fig. 4; Supplementary Tables 7–8). Logistic and Poisson regression were used for dn variants, and logistic and negative binomial regression for rare variants. Analyses of dn and transmitted variants were conducted separately for individuals with OCD, CTD, and a combined OCD and CTD (OCDCTD) cohort. Joint analysis was motivated by evidence of significant overlap in dnDam variants between OCD and CTD (Fig. 2a; Supplementary Table 9). Rare variant analyses were restricted to the OCD cohort due to the limited number of available CTD singleton samples. To mitigate batch effects due to differences in sequencing coverage and exome capture platforms, all analyses were limited to high-confidence consensus callable coding regions. Statistical models were implemented in R, and 95% confidence intervals were calculated as the estimate ± 1.96 × standard error (SE). Full model specifications are provided below:

- Dn variant, logistic regression: *case status ∼ variant count + consensus callable base pairs*
- Dn variant, Poisson regression: *variant count ∼ case status + offset(log(consensus callable base pairs))*
- Transmitted rare variant, logistic regression: *case status ∼ transmitted variant count + untransmitted variant count + sex + ancestry PCs 1-10*
- Transmitted rare variant, negative binomial regression: *transmitted variant count ∼ case status + untransmitted variant count + sex + ancestry PCs 1-10*
- Rare variant, logistic regression: *case status ∼ variant count + sex + ancestry PCs 1-10*
- Rare variant, negative binomial regression: *variant count ∼ case status + sex + ancestry PCs 1-10*

These complementary models allowed us to evaluate both the frequency of variant carriers and the overall burden per individual, with consistent findings across approaches. We observed an exome-wide increased burden in cases versus controls for damaging dn and for rare variants, but not for transmitted variants; therefore, transmitted variants were excluded from gene discovery analyses (see Supplementary Note 2 for further details on variant class selection).

To assess potential sex differences in dn burden, we examined variant rates stratified by sex across all cohorts (OCD, CTD, OCDCTD). Among trios, the proportion of male probands varied by cohort (CTD: 79%; OCD: 59%; controls: 47%; Supplementary Table 11), consistent with the known male sex bias in CTD^6^. Within each cohort, we used Poisson regression to compare dn variant rates between male cases and male controls, female cases and female controls, and male cases versus female cases (Supplementary Fig. 8; Supplementary Table 12). No consistent or statistically significant sex differences were observed.

#### Contribution of rare variants to risk

To estimate the proportion of observed variants that contribute to disease risk, we calculated the difference in variant rates between cases and controls, then divided by the observed rate in cases. Rate ratios (RR) and 95% confidence intervals were estimated using Poisson regression (for dn variants from family-based data) or negative binomial regression (for rare inherited variants from singleton data), incorporating the offsets and covariates described above. The proportion of risk-mediating variants was calculated as 1 - 1/RR, with 95% confidence intervals derived from the log-transformed bounds of the RR estimates (Supplementary Table 14).

To estimate the percentage of probands carrying at least one risk-contributing variant, we compared the percentage of individuals with at least one qualifying variant in case versus control cohorts. For family-based data, we considered individuals carrying at least one dnDam (dnLGD or dnMis3) variant. For singleton data, we considered individuals with at least one rare LGD in genes with LOEUF in the 0– 20% range. The difference in carrier proportions between cases and controls was used as a proxy for the proportion of individuals in which a variant of that class contributes to disease risk. The 95% confidence interval was estimated using bootstrapping (1,000 replicates) (Supplementary Table 15).

### Cross-disorder overlap of *de novo* damaging variants

We tested whether dnDam genes were shared more than expected by chance between OCD and CTD subgroups using family-based cohorts with balanced sizes: OCDwCTD, CTDonly, and OCDonly/unk (OCDonly and OCDunk combined due to the small cohort size of OCDonly). Neurotypical SSC siblings served as a negative control. For each subgroup, we counted the number of unique genes with dnLGD or dnMis3 variants, then generated 10,000 random gene sets matched in size and weighted by mutability to simulate the null expectation. We quantified observed gene overlaps between cohort pairs and compared them to the null distributions to calculate one-sided empirical *p*-values and *z*-scores.

Benjamini-Hochberg correction was applied for multiple testing (Fig. 2a; Supplementary Table 9).

### Gene discovery

#### Estimating the number of underlying risk genes

The number of risk genes contributing to CTD, to OCD, and to OCD/CTD was estimated using two distinct methods: 1) a previously established maximum likelihood estimation (MLE) procedure^22,23,97^ and 2) a framework developed for the “unseen” species problem^22,98^. All variants in the “restricted callable” interval described above were included in the analysis. Both approaches gave similar estimates (Supplementary Table 13); MLE values were used for downstream analyses, consistent with prior studies^22–24^.

#### MLE

For each cohort, we simulated 10,000 permutations for candidate risk gene counts from 1–3,000, matching the observed number of dnDam variants. In each simulation, a proportion of variants (equal to the estimated fraction carrying risk) was randomly assigned to risk genes, and the remainder to non-risk genes, weighted by gene-level mutability. We compared the simulated and observed distributions of genes with 1, 2, or ≥3 damaging variants, calculated the likelihood of concordance, and smoothed results using a sliding window of 10 genes. The median number of genes within the window with the highest mean likelihood was taken as the final MLE estimate.

#### Unseen species

The total number of risk genes (*C*) was also estimated using the “unseen species” framework^22,98^:

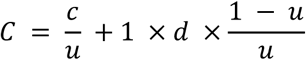

where:

- d = number of risk-carrying variants = observed damaging variants – expected damaging variants (expected values scaled from SSC control trios)
- c = observed risk genes = d – recurrent variants + genes with recurrent variants.
- c₁ = genes mutated once = c – genes with recurrent variants.
- u = probability that a new variant hits a previously mutated gene = 1 – (c₁/d).

Confidence intervals were obtained by recalculating C using Poisson-derived bounds for expected control dnDam counts, as previously described^24^. Estimates were generated for OCD, CTD, and OCDCTD using dn variants.

#### TADA framework for gene discovery

We applied the Transmission and *De Novo* Association (TADA) test, a Bayesian model that integrates evidence across multiple classes of genetic variation to identify risk genes ^42^. We used an updated version of TADA that accommodates both dn and rare inherited variants, allows for flexible inclusion of gene-level constraint metrics and functional annotations, and takes into consideration variants observed in controls^30^ (TADA R code from: https://github.com/talkowski-lab/TADA_2022; https://doi.org/10.5281/ zenodo.6496480). TADA computes a Bayes factor (BF) for each gene and variant class based on observed variant counts, cohort sizes, estimated number of underlying risk genes, and gene- and variant class-specific prior probabilities. Gene-level BFs are obtained by multiplying BFs across variant classes, then converted into false discovery rate (FDR) q-values to identify risk genes. For primary gene discovery, we included variant classes that showed significant enrichment in burden analyses:

- OCD and OCDCTD cohorts: dnLGD (stratified by LOEUF 0–40% vs. 40–100%), dnMis3, and rareLGD in LoF-intolerant genes (LOEUF 0–20%) from singleton cases.
- CTD cohort: dnLGD (stratified by LOEUF 0–40% vs. 40–100%) and dnMis3.

TADA requires the following inputs per gene and variant class:

- **Sample sizes:** number of trios (for dn variants) or singletons (for rare inherited variants).
- ***π* (pi):** the fraction of causal genes, estimated as the MLE-based number of risk genes divided by the total number of genes in the coding consensus region (n = 18,705).
- **ì (mu)**: mutability, or gene-level mutation rate for each variant class, derived from gnomAD v4.1 gene constraint metrics. LGD mutability was estimated by lof.mu, the sum of mutation rates across all possible high-confidence pLoF variants in a transcript. Missense mutability was estimated by mis.mu. Subcategory-specific missense mutability was estimated by scaling mis.mu by the observed proportion of each missense subcategory in control samples. For example, Mis3 mutability was obtained by multiplying mis.mu by the fraction of Mis3 variants among all missense variants observed in SSC control individuals within the singleton consensus interval. TADA analysis included only the 18,111 genes with available constraint data, resulting in the exclusion of a small number of variants (for example, 8 of 403 dnLGD variants).
- **ë (lambda):** fold-enrichment of variants in cases vs. controls, derived from burden analyses using Poisson regression for dn variants from trios and negative binomial regression for rare variants from singletons.
- Ɣ **(gamma):** relative risk for each variant class, calculated as:

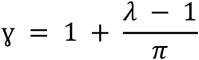

Relative risks were floored at 1 to exclude estimates suggesting protective effects, which were not considered in this analysis.

The specific pi, mu, and gamma values used in TADA are provided in Supplementary Table 16. TADA was used to compute Bayes factors (BF) for each gene within each variant class. The total gene-level BF was calculated by multiplying BFs across all variant classes. All original BF values (including those less than 1, providing evidence against association) were retained in gene discovery analyses, reflecting a more conservative approach than studies that floor BFs at 1^30^ (discussed in Supplementary Note 3). BFs were then converted to posterior probabilities and used to estimate false discovery rate (FDR) *q* values. Previously established statistical thresholds were applied to define high-confidence (hc, FDR < 0.1) and probable (p, 0.1 ≤ FDR < 0.3) risk genes^22–24,43^. To reduce the likelihood of spurious signals —particularly in smaller cohorts (e.g., CTD and OCD analyses) and genes with low expected mutability, where some p genes were supported by only a single dn observation—we elected to require p risk genes to have at least two dnDam variants. TADA was run separately for the OCD, CTD, and combined OCDCTD cohorts (Fig. 2; Supplementary Fig. 9; Supplementary Table 16). For the OCD and OCDCTD cohorts, analyses were performed both with trios only and with trios and singletons to assess the impact of adding singleton data (Supplementary Table 16, Supplementary Note 3). The number of hc and p genes identified in these analyses are reported in Supplementary Table 17.

### Risk gene characteristics

#### Risk gene architecture

We estimated relative risk (RR) across three classes of genetic variation: dnDam variants, rare LGD variants, and common variants. Each gene was assigned a single RR estimate per cohort (OCD, CTD, OCDCTD) based on case versus control variant rates, prioritizing variant classes in the following order: dn > rare > common. For dn variants, RR was defined as the observed number of dnDam variants in probands divided by the expected number of dn mutations per gene, based on gene-specific loss-of-function and missense mutation rates from gnomAD v4.1 (e.g. TADA μ). For rare variants, RR was calculated as the ratio of allele frequencies in OCD singleton probands (n = 1,546) versus controls from the gnomAD v4.1 exome dataset (n = 730,947). For common variants, we used odds ratios and allele frequencies reported for genome-wide significant loci in a recent OCD GWAS meta-analysis^13^. Only one gene (*CCDC168*) was included based on rare LGD burden alone, with no dnDam variants observed in OCD or OCDCTD probands. A LOESS-smoothed trend line with 95% confidence interval was plotted to visualize the overall pattern of RR across the population allele frequency spectrum (Fig. 2; Supplementary Fig. 10; Supplementary Table 18).

#### Variant-class and cohort contributions to risk gene associations

To examine the genetic association contributions for OCD, CTD, and jointly implicated risk genes, we focused on a set of 36 genes that reached hc status in OCD, CTD, and/or combined OCD+CTD analyses. For each of these genes, we assessed the sources of genetic evidence across all available OCD and CTD data. Gene-level association evidence was computed using the TADA framework, which integrates BFs from three variant classes: dnLGD, dnMis3, and rare LGD. To understand the sources of this evidence, we quantified the relative contribution from: (1) sample phenotypes (e.g. OCD and CTD status, including OCDonly, OCDunk, OCDwCTD, CTDonly, CTDunk); and (2) variant class (e.g., dnLGD, dnMis3, rareLGD). For (1), we calculated a phenotype-specific contribution score for each gene and variant class:

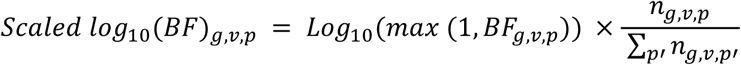

- *BF_g,v,p_* is the Bayes factor for gene *g,* variant class *v,* and phenotype *p*.
- *n_g,v,p_* is the number of variants from samples with phenotype *p*.
- *p′* indexes all phenotype groups (e.g., OCDonly, OCDunk, OCDwCTD, CTDonly, CTDunk).

Contributions were normalized across cohorts for each gene and variant class. We also computed overall Bayes factors for OCD (combining OCDonly, OCDunk, and OCDwCTD) and for CTD (combining CTDonly, CTDunk, and OCDwCTD). For visualization, BFs below 1 were floored at 1, which affected three genes (*WWC1, CESLR3, ABCD2;* discussed in Supplementary Note 3). Cohort-specific contributions were then summed across variant classes to yield the total support from each diagnostic group. This decomposition allowed us to evaluate how genetic association evidence was distributed across variant types and phenotypes (Fig. 3; Supplementary Fig. 11; Supplementary Table 19).

### Risk gene overlap

#### Overlap with external risk genes

We evaluated overlap between OCD and related gene sets (hcOCD, hcCTD, hcOCDCTD) and risk gene sets identified from rare variant-based WES studies in autism spectrum disorder (ASD), developmental disorders and intellectual disability (DD/ID), neurodevelopmental disorders (NDD), schizophrenia (SCZ), congenital heart defects (CHD), and genome-wide association studies (GWAS) of OCD and SCZ. We defined the following gene sets from rare variant studies

- SFARI Tier 1S (ASD): SFARI Tier 1 and syndromic genes (release 01-13-2025) downloaded from SFARI Gene^28^.
- Zhou 2022 curated (ASD/NDD): set of curated dominant or X-linked ASD or NDD genes reported in Zhou et al. (2022)^32^.
- Satterstrom 2020 (ASD): genes with TADA FDR < 0.1^29^.
- Fu 2022 (ASD): genes with TADA FDR < 0.1^30^.
- Trost 2022 (ASD): genes from MSSNG with TADA+ FDR<0.1^31^.
- Zhou 2022 (ASD): genes from SPARK that met study-wide significance^32^.
- Kaplanis 2020 (DD/ID): severe developmental disorder risk genes that were exome-wide significant^33^.
- Singh 2022 (SCZ): genes from SCHEMA with meta-analysis q < 0.1^34^.

We defined the following gene sets from common variant (GWAS) studies

- Strom 2025 - functional (OCD): a functionally mapped gene set defined by Strom et al. (2025)^13^, consisting of genes linked to 30 lead SNPs from the primary OCD GWAS meta-analysis through at least one of six gene-based mapping approaches. These genes were extracted from Strom et al. 2025 Table Supplementary Table 14, and the original 251 genes were filtered to 243 genes after mapping to HGNC symbols (8 genes without matching HGNC symbols).
- Strom 2025 - positional (OCD): a positionally mapped gene set consisting of n = 69 genes located within ±50 kb of 61 lead SNPs identified in the OCD GWAS meta-analysis^13^. These 61 SNPs reached genome-wide significance either in the full sample (main GWAS) or in analyses of four subgroups defined by ascertainment: cases identified by a health care professional in a clinical setting, from health records or biobanks, ascertained for a different psychiatric disorder, or by self-reported diagnosis in the 23andMe consumer-based setting. The SNPs were extracted from Supplementary Table 3 of Strom et al. (2025).
- Trubetskoy 2022 (SCZ): gene set prioritized by Trubetskoy et al. (2022)^82^, consisting of 106 protein-coding genes (only 105 with HGNC symbols) associated with 287 lead schizophrenia SNPs through fine-mapping and functional genomic data.

We evaluated the overlap between hcOCD, hcCTD, and hcOCDCTD gene sets with each of the above risk gene sets (Fig. 3; Supplementary Fig. 12). We conducted two-sided Fisher’s exact tests (FET) to evaluate the pairwise overlap significance. The background for each comparison was defined as the intersection of genes analyzed in both studies, when available. We constructed 2×2 contingency tables based on whether genes were present in the two sets or not. Observed *p*-values from FETs were corrected for multiple testing using the Benjamini–Hochberg false discovery rate (FDR). To assess empirical significance, we performed 10,000 mutability-weighted permutations. In each iteration, we randomly sampled gene sets matched in size to the hcOCD, hcCTD, or hcOCDCTD sets, with sampling probability based on gene-level loss-of-function (LoF) mutability from gnomAD v4 (see TADA mu). For comparisons involving whole-exome sequencing (WES) data, the background was further restricted to genes evaluated in the respective study. Empirical p-values were calculated as the proportion of permutations with odds ratios greater than or equal to the observed value (Fig. 3; Supplementary Table 20).

#### Enrichment for OCD common variant risk

Gene-level *z*-scores were computed using MAGMA v1.10^44^ from OCD GWAS^13^ meta-analysis summary statistics. Summary statistics were downloaded from https://pgc.unc.edu/for-researchers/download-results/, and were generated from 23,493 European ancestry cases and 1,114,613 controls (note: this excluded 23andMe samples, which comprised over half of the case cohort in Strom et al. 2025). SNPs were mapped to genes using GENCODE v47 (GRCh37), with the default ‘snp-wise = mean’ model. Linkage disequilibrium was accounted for using 1000 Genomes European reference data. Gene-level results are visualized in Supplementary Fig. 12 and discussed in Supplementary Note 5.

#### Cross-disorder dnLGD overlap

We assessed whether dnLGD variants were shared more than expected by chance between pairs of OCDCTD, ASD^30^, DD/ID^33^, and SCZ^34^. dnLGD from unaffected SSC siblings were used as negative controls for all comparisons. We focused on dnLGD because they were consistently reported across all WES studies. Observed gene overlap, null distributions, *z*-scores, and Benjamini-Hochberg p-value corrections were computed as above in *Cross-disorder overlap of* de novo *damaging variants* (Fig. 3e, Supplementary Fig.13; Supplementary Table 21). To assess the magnitude of gene overlap, we performed one-sided Fisher’s exact tests on genes affected by dnLGD, generating odds ratios (OR) and 95% confidence intervals for each cohort pair. The background universe was restricted to 18,111 genes evaluated in TADA, and p-values were corrected for multiple hypothesis testing using the Benjamini–Hochberg method (Supplementary Table 21). Interpretation for SCZ is limited due to the very small gene set.

#### Comparison of gene-level association across disorders

To compare gene-level association strength across disorders, we compiled q-values from recent large-scale WES studies of OCD and CTD (this study), ASD, NDD, and SCZ^30,33,34^. Q-values were transformed as –log₁₀(q), capped at 10⁻⁸, and min– max scaled within each disorder to allow for cross-disorder comparison. Scaled values were visualized using ternary plots to illustrate the relative strength of association for each gene across sets of three disorders (see Supplementary Note 6; Supplementary Fig. 14, Supplementary Table 22).

### Region, age, and cell specific expression of risk genes

#### GTEx tissue-specific gene expression enrichment analysis

Tissue-specific expression of hcOCDCTD genes was assessed using bulk RNA-seq data from 54 tissues in the Genotype Tissue Expression Project (GTEx; v10)^46^. Expression values were normalized as log_10_(Transcripts Per Million [TPM] + 1). Gene expression specificity for each tissue was quantified by calculating t-statistics comparing expression in the focal tissue against others, excluding related tissues^53^. The background gene set was defined as the intersection of all genes analyzed in both GTEx and this study (n = 17,970). Gene set enrichment analysis (GSEA) was performed for each tissue using the R package fgsea v1.26.0. For each tissue, genes were ranked by *t*-statistic in descending order, and enrichment of OCDCTD risk genes was tested using a one-sided enrichment model (scoreType = “pos”). *P*-values were corrected for multiple testing using the Benjamini–Hochberg method (Fig. 4; Supplementary Table 23).

#### Brain spatiotemporal enrichment analysis

We evaluated temporal expression patterns of risk gene sets using expression data from the BrainSpan Atlas of the Developing Human Brain (www.brainspan.org)^47,48^, which profiled gene expression across multiple brain regions and developmental stages. Samples were grouped by anatomical region, focusing on the neocortex, striatum, thalamus, and cerebellum due to sample size considerations, and analyses were conducted separately for each region. Within each region, samples were grouped into four developmental stages: prenatal (before birth), infancy/childhood (birth–12 years), adolescence (12–20 years), and adulthood (≥20 years). Normalized gene expression values were scaled across samples to compute temporal specificity *z*-scores, with higher values indicating greater stage-specific expression. For each developmental stage, the mean *z*-score per gene set provided a measure of relative stage-specific expression.

Disease-associated gene sets were compared to a background set (genes evaluated by TADA or WES and present in BrainSpan) using a one-sided Wilcoxon rank-sum test, with *p*-values corrected for multiple testing via the Benjamini–Hochberg method across 60 comparisons (15 gene sets × 4 stages). ASD genes were included as a positive control for mid-fetal cortical expression bias^29,30^, and Alzheimer’s disease (AD)-associated genes^54^ as a positive control for late-adulthood expression bias. HcASD genes were defined as above, and 76 AD genes were obtained from Bellenguez *et al*. 2022 Supplementary Table 5^54^. For all analyses, findings were consistent between the BrainSpan microarray dataset^48^, which has a larger sample size, and the RNA-seq dataset^47^ (Fig. 4; Supplementary Fig. 15; Supplementary Table 24).

We conducted the same analysis with a dataset of rhesus macaque brains spanning early prenatal through adult development^49^. This non-human primate dataset served as a comparator, offering lower variability in sample collection and postmortem quality compared with human brain atlases. Findings were broadly consistent with those obtained from human data (Fig. 4; Supplementary Table 24).

#### Enrichment in Zeisel 2018 (scRNAseq of mouse nervous system)

We tested whether OCD- and CTD-associated risk genes were enriched in cell type–specific markers from a large-scale single-cell RNA-seq study of the mouse nervous system^50^. Marker genes for 39 broad cell types (level 4) were defined by Bryois et al. (2020)^55^ as the top 10% most specific genes based on expression specificity scores (i.e., proportion of a gene’s expression in each cell type relative to all cell types). Marker sets were obtained from Supplementary Table 15 of Bryois et al.^55^. We used one-sided Fisher’s exact tests (FETs) to test enrichment of hc risk gene sets (hcOCD, hcCTD, hcOCDCTD) against each cell type’s marker set. The background gene set was the intersection of all tested genes and those in Zeisel et al. (n = 17,970). P-values were corrected using the Benjamini–Hochberg method across all cell types and risk gene sets (Fig. 4; Supplementary Fig. 16; Supplementary Table 25).

#### Enrichment in Xu 2014 (BAC-TRAP of mouse CNS cell types)

To assess enrichment using an orthogonal approach, we analyzed brain cell type–specific markers from BAC–TRAP data^51,83^, which measures ribosome-bound mRNA in genetically defined mouse CNS cell types. Gene specificity was quantified using the specificity index (pSI), a rank-based score with empirical p-values from permutations. We defined cell type markers as genes with pSI < 0.05, downloaded from the pSI and pSI.data R packages (https://sites.wustl.edu/doughertylab/psi_package-page/). Human orthologs were mapped using Ensembl v75 and converted to HGNC symbols. Background gene sets consisted of all genes that passed the original expression and quality control filters. Risk gene enrichment was tested via one-sided FETs. *P*-values were corrected using the Benjamini–Hochberg method (Fig. 4; Supplementary Fig. 16; Supplementary Table 26).

#### Enrichment in Siletti 2023 (snRNAseq of human brain)

We also tested for enrichment in human brain cell types using snRNAseq data from Siletti et al. (2023)^52^, which profiled ∼3 million nuclei and defined 31 supercluster cell types. Gene expression specificity was computed as each gene’s TPM in a given cell type divided by its total TPM across all types^55^. GSEA was performed for each supercluster and risk gene set as described above for GTEx. P-values were adjusted using the Benjamini–Hochberg method across all cell types and gene sets (31 superclusters × 3 gene sets) (Fig. 4; Supplementary Fig. 16; Supplementary Table 27).

### Gene set enrichment analysis

We performed gene set overrepresentation analysis using NetworkAnalyst^56^. To balance statistical power with biological relevance, we focused on the union of hcOCDCTD and pOCDCTD risk genes, representing genes likely to be functionally connected and enriched for true disease risk. Enrichment was conducted for Gene Ontology Biological Process (GO:BP) terms using default parameters, which apply multiple hypothesis correction with the Benjamini-Hochberg method (Supplementary Fig. 17; Supplementary Table 28).

### Risk gene connectivity

To evaluate the molecular network connectivity of risk gene sets, we evaluated hc and p gene sets across two complementary gene interaction networks: BioGRID and PCNet^57,59^. These two networks provide alternative approaches to determining gene connectivity—BioGRID captures physical protein-protein interactions (PPIs) of the proteins encoded by risk genes, and PCNet captures broader functional gene interactions across multiple interaction types (including PPIs). BioGRID v4.4.241 was restricted to multi-validated physical interactions, as defined by BioGRID (https://wiki.thebiogrid.org/doku.php/biogrid_mv). PCNet v2.2 (which excludes co-citation-derived edges) was downloaded from the Network Data Exchange (NDEx^99^, www.ndexbio.org). All network gene identifiers were updated to current HUGO (Human Genome Organization) Genome Nomenclature Committee (HGNC) gene symbols.

#### Within-set connectivity

Connectivity was measured as the number of risk genes directly connected to another gene in the same risk gene set (n_connected). Significance was assessed using 10,000 permutations of random gene sets matched for size and drawn from the network-specific background. Empirical one-sided *p*-values were calculated as the fraction of permutations with n_connected ≥ observed, and *z*-scores were computed relative to the empirical null (Fig. 5; Supplementary Fig. 18; Supplementary Table 29).

#### Interactome overlap

To assess network-level coherence, we tested whether pairs of risk genes shared more interactors than expected by chance. For each unique pair within a gene set, we performed a one-sided Fisher’s exact test for shared interactors; the background universe was defined as all TADA genes that were represented in the network. The proportion of significant gene pairs (p < 0.05) was then compared to an empirical null distribution generated from 1,000 samples of 100 randomly drawn gene pairs. *Z*-scores were computed to evaluate deviation from the empirical null Fig. 5; Supplementary Fig. 18; Supplementary Table 30).

### Rare and common variant convergence in interactomes

We evaluated whether genes that interact with OCD and CTD risk genes are enriched for genes carrying dnLGD variants in trio probands or for genes implicated by OCD common variant association^19^. hcOCD, hcCTD, and hcOCDCTD were used as “seed genes” to identify first-degree interactors in two large-scale gene interaction networks, BioGRID and PCNet (described above)^57,59^. Seed genes were excluded from being interactors. The background gene universe was defined as the intersection of genes present in the network and those evaluated in this study (TADA).

#### Interactome enrichment for dnLGD

To test whether the first-degree network interactors of hc and p risk genes were themselves enriched for rare deleterious variants, we evaluated the burden of dnLGD relative to dnSyn observed in trio probands using Fisher’s exact tests (two-sided), with Bonferroni correction for multiple hypothesis testing. As above, we performed 10,000 permutations, generating matched random gene sets from the network background to evaluate empirical p-values. A result was considered significant if it met both criteria: Bonferroni-corrected Fisher p-value and empirical p-value < 0.05 (Fig. 5, Supplementary Table 31).

#### Interactome enrichment for common variant risk

To assess whether common variant-associated OCD genes were overrepresented among first-degree interactors of rare variant risk genes, we tested for enrichment of genes mapped to OCD GWAS loci (either positionally or functionally, described in *Overlap with external risk genes*). We performed two-sided Fisher’s exact tests using 2×2 contingency tables comparing interactor status (yes/no) with common variant association status (yes/no), with Bonferroni correction for multiple hypothesis testing across all tested gene set x network combinations. To validate the observed enrichments, we performed permutation testing: for each rare variant gene set, we generated 10,000 random gene sets of matched size from the network-specific background. For each, first-degree interactors were identified, and enrichment for common variant-associated genes was recalculated using Fisher’s exact test. Empirical p-values were defined as the proportion of permuted sets with odds ratios greater than the observed value. Enrichment was considered significant if both the Bonferroni-adjusted Fisher p-value and the empirical p-value were < 0.05 (Fig. 5, Supplementary Table 32). We additionally tested the association of OCD common variant risk^19^ on first-degree interactors using MAGMA gene set analysis (GSA)^44^(Supplementary Fig. 18; Supplementary Table 33). We applied competitive GSA with standard covariates, which account for gene size, within-gene LD, inverse-mean minor allele count, and effective sample size per SNP.

### Network propagation and functional enrichment

We evaluated the functional roles of hc risk genes using network propagation, which distributes signals from a set of ‘seed’ risk genes across the PCNet v2.2 interaction network to quantify how functionally connected each gene is to the seed set. Analyses were performed separately for hcOCD (n = 12), hcCTD (n = 10), and the union of hcOCD/hcCTD/hcOCDCTD (n = 36) seeds. Network propagation was implemented using a random walk with restart approach (restart probability = 0.1) as previously described^100^, with seed genes assigned a starting value of 1 and all other PCNet genes assigned 0. To assess significance, we performed a permutation test in which initial gene labels were randomly shuffled 20,000 times. Gene-level empirical *p*-values were calculated as the fraction of random propagations in which propagated scores were greater than or equal to the observed scores. The top 100 genes with the lowest permutation-based p-values (indicating the strongest connectivity to the seed set) were selected for overrepresentation analysis (ORA). ORA was conducted using Gene Ontology: Biological Processes (GO:BP v7.5.1) with R clusterProfiler::enricher (minGSSize = 10, maxGSSize = 500 (default), pAdjustMethod = “fdr”, universe = all PCNet genes in GO:BP, pvalueCutoff and qvalueCutoff = 1.1 to retain all terms for post hoc simplification and clustering). To reduce redundancy among enriched GO terms, terms with overlapping gene membership were clustered based on Jaccard similarity, and terms with fully overlapping genes were collapsed. Within each cluster, the representative GO term was chosen as the term with the lowest adjusted p-value. From these representative terms, the top 10 most significant terms per gene set were retained for visualization. Heatmaps were visualized using R ComplexHeatmap (Supplementary Fig. 19; Supplementary Table 34).

### Use of generative AI

Portions of the methods were drafted, and the main text refined, with assistance from ChatGPT (OpenAI, GPT-4o and GPT-5). All content was reviewed, edited, and approved by the authors, who take full responsibility for the final manuscript.

