## Supplementary Information for "Rare coding mutations identify 36 large-effect risk genes in obsessive-compulsive disorder and chronic tic disorders"

### Table of Contents:

Supplementary Figures 2
Supplementary Table Legends 21
Supplementary Notes 22
Supplementary Note 1: Study participants 22
Supplementary Note 2: Rationale for variant class selection for gene discovery 25
Supplementary Note 3: Interpretation of high-confidence genes with atypical rare LGD patterns and those unique to the CTD cohort 27
Supplementary Note 4: Comparison with previously reported risk genes 29
Supplementary Note 5: Gene-level MAGMA analysis of OCD GWAS 30
Supplementary Note 6: Comparison of gene-level association across disorders 31

### Supplementary Figures

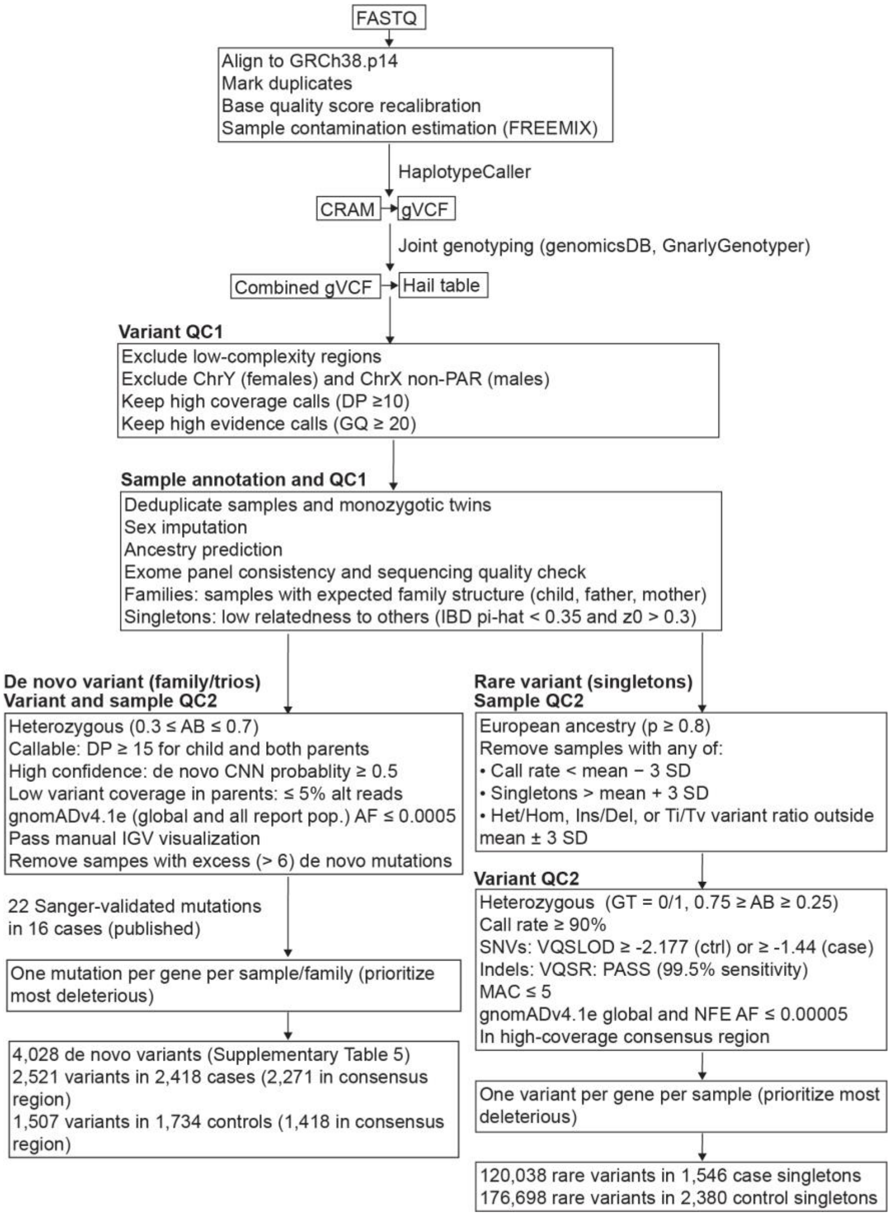


**Supplementary Figure 1 | Sample and variant data processing overview**. Overview of sample, genotype, and variant quality control for whole-exome sequencing data. Raw sequencing reads (FASTQ) were aligned to GRCh38 and processed with the Genome Analysis Toolkit (GATK) for base recalibration and joint genotyping, yielding raw variant calls (VCF). Joint-genotyped VCFs underwent quality control to define dn (family-based) and rare (singleton) variant sets for downstream analyses. Low-complexity regions were defined as previously described[84]. Variant quality score recalibration (VQSR), chimeric read estimation, and contamination metrics were computed using GATK. Relatedness and sample contamination were evaluated with VerifyBamID2. Common variants were excluded based on allele frequency in gnomAD v4.1 exomes. Transmitted variants, not included in gene discovery, are not shown. Abbreviations: AB, allele balance; AF, allele frequency; Chr, chromosome; CRAM, compressed reference-based alignment format; DP, read depth; FASTQ, raw sequencing read format; GT, genotype; GQ, genotype quality; GRCh38: Genome Reference Consortium Human Build 38; gVCF, genomic variant call format (includes non-variant sites); gnomAD v4.1e, genome aggregation database version 4.1 exome dataset; IBD, identity by descent; IGV, Integrative Genomics Viewer; NFE, non-Finnish European; PAR, pseudoautosomal region; QC, quality control; SD, standard deviation; VQSLOD: variant quality score log-odds; VQSR, variant quality score recalibration.

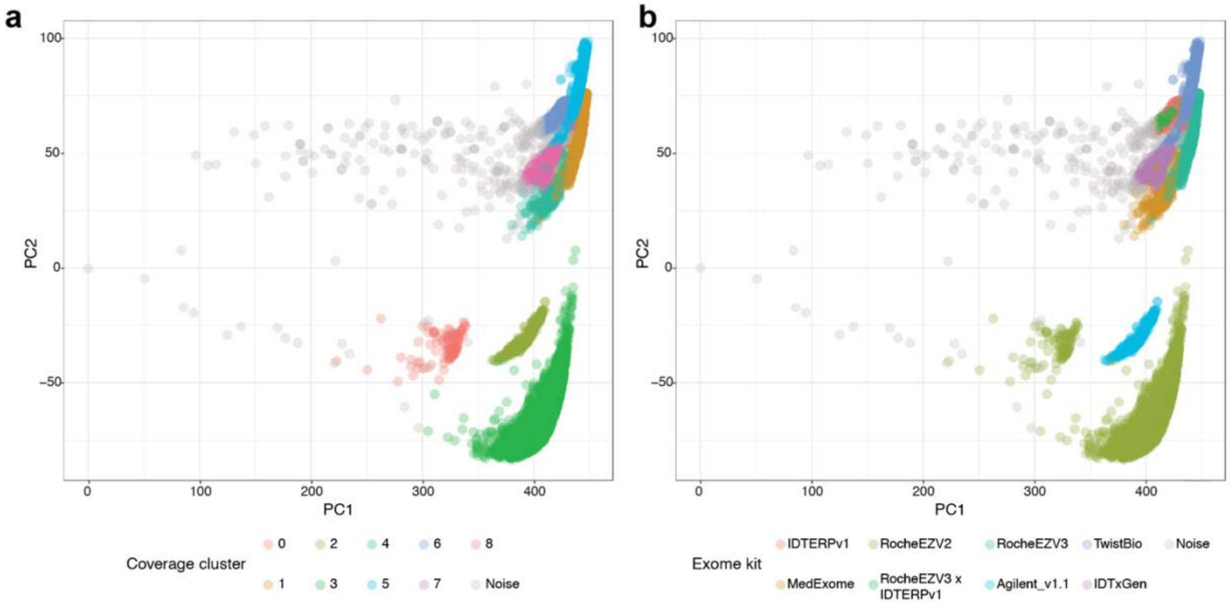


**Supplementary Figure 2 | Exome capture platform inference. a,** Clusters identified by HBDSCAN (hierarchical density-based spatial clustering of applications with noise); unclustered samples are labeled as noise. **b,** As in a, but colored by annotated exome capture kit: Roche SeqCap EZ Exome v2 (RocheEZV2), Roche SeqCap EZ Exome v3 (RocheEZV3), Twist Bioscience Comprehensive Exome Panel (TwistBio), Roche SeqCap EZ MedExome (MedExome), Agilent SureSelect v1.1 (Agilent_v1.1), IDT Exome Research Panel v1 (IDTERPv1), IDT xGen Exome Research Panel (IDTxGen), and RocheEZV3 or IDTERPv1 where capture kit was unspecified in publications (RocheEZV3 × IDTERPv1).

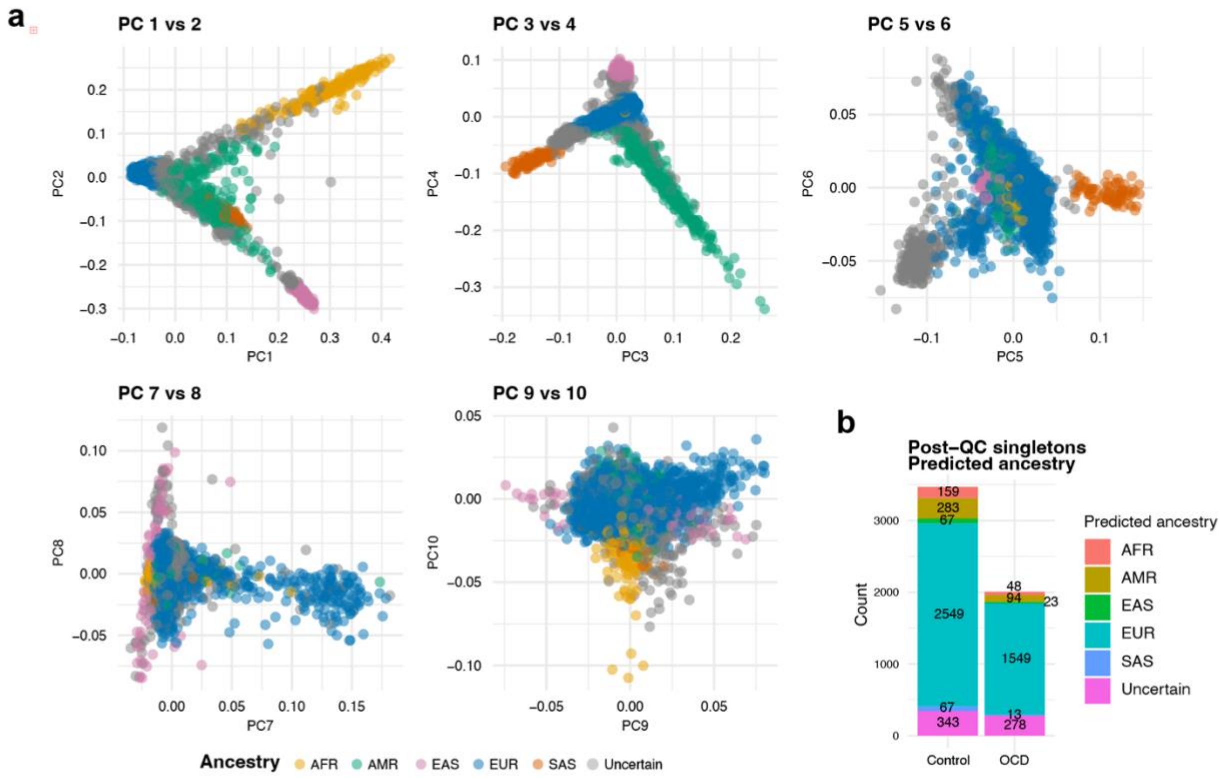


**Supplementary Figure 3 | Ancestry assignment and exome capture of singleton samples. a**, Ancestry of 5,473 singleton samples after the first round of sample quality control (QC), before removing non-EUR samples. The first 10 ancestry principal components are shown. Global ancestry was inferred using a random forest model trained on 1000 Genomes individuals. **b**, Distribution of ancestry assignments for post-QC singleton samples. AFR, African/African American; AMR, Admixed American; EAS, East Asian; EUR, European; SAS, South Asian.

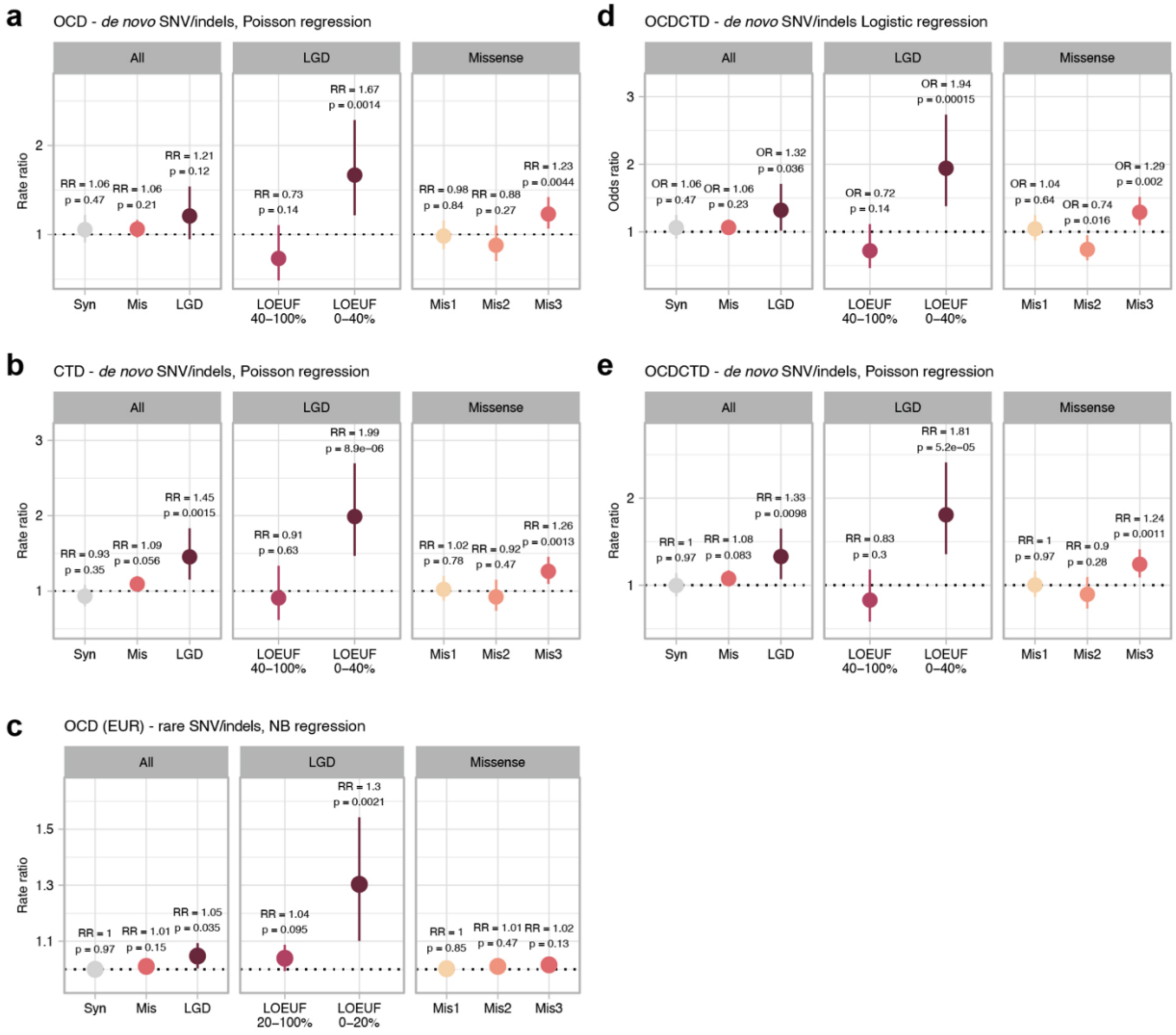


**Supplementary Figure 4 | Burden testing of *de novo* and rare variants. a,b,e**, Poisson regression comparing dn variant rates in OCD (**a**), CTD (**b**), and OCDCTD (**e**) families versus controls. **c,** Negative binomial regression comparing rare variant burden rates in OCD singletons versus controls. **d**, Case-level logistic regression testing for association between rare variants and case status in OCDCTD. Rate ratios (Poisson/negative binomial) and odds ratios (logistic) are shown with 95% confidence intervals. Consistent with established convention[22–24,30,34,37], *p*-values are not corrected for multiple tests. Allele frequency thresholds in a population reference[36] were ≤ 0.05% for dn variants and ≤ 0.005% for singleton rare variants (see Methods). Results are shown for (left panels) "All" variants (synonymous, missense, and likely gene-disruptive [LGD]), (middle panels) "LGD" variants stratified by genic constraint (lower LOEUF deciles indicate higher constraint), and (right panels) "Missense" variants classified by predicted deleteriousness (Mis3 = most deleterious[38]).

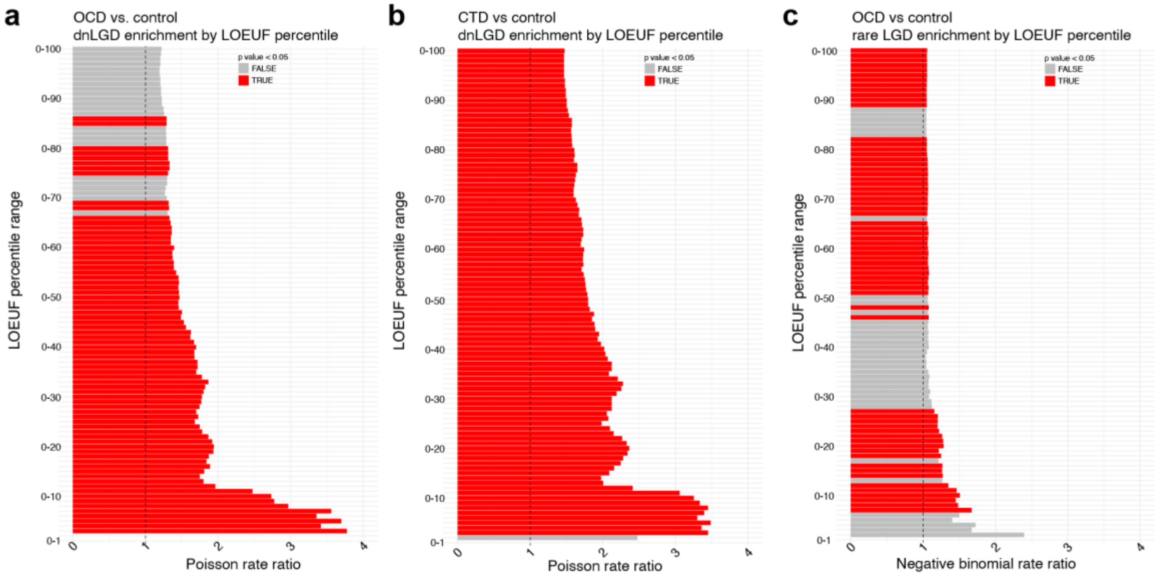


**Supplementary Figure 5| Selection of LOEUF cutoffs for LGD variant burden analyses.** Burden analyses of dnLGD variants in OCD (**a**) and CTD (**b**), and rare LGD variants in OCD (**c**), stratified by LOEUF. Genes were ranked by LOEUF percentile, and cumulative bins were assessed from the most LoF-intolerant (0–1%) to all genes (0–100%) in 1% increments. LGD counts were modeled as a function of disease status using Poisson regression for dn variants (**a**, **b**) and negative binomial regression for rare variants (**c**). Red color indicates significance ($p$ < 0.05, uncorrected for multiple testing). Dashed line indicates rate ratio = 1. LOEUF cutoffs of 40% and 20% were selected for dn and rare variants, respectively.

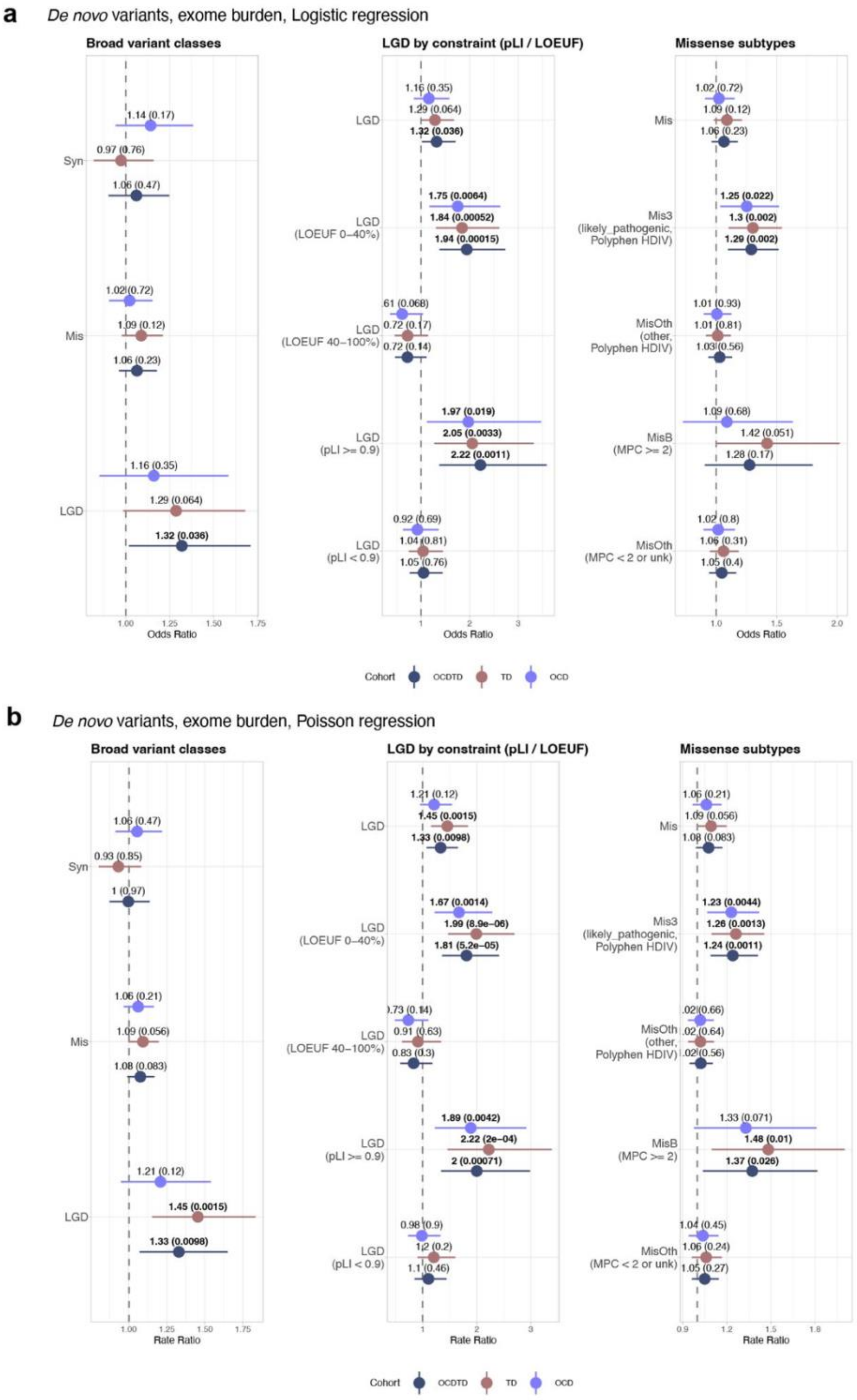


**Supplementary Figure 6 | *De novo* variant enrichment in OCD, CTD, and OCDCTD cohorts, stratified by genic constraint and missense deleteriousness. a**, Logistic regression models testing association between dn variant and case status. **b**, Poisson regression comparing dn variant rates in cases versus controls. Left: broad variant classes (synonymous, missense, and likely gene-disruptive [LGD]). Middle: LGD stratified by genic constraint (pLI, LOEUF; lower LOEUF/higher pLI = greater constraint). Right: missense variants stratified by predicted deleteriousness (Mis3 from PolyPhen-2 HDIV, and MisB from MPC). Odds ratios (ORs; logistic regression) and rate ratios (RRs; Poisson regression) are shown with 95% confidence intervals. *p* values are two-sided and uncorrected for multiple testing.

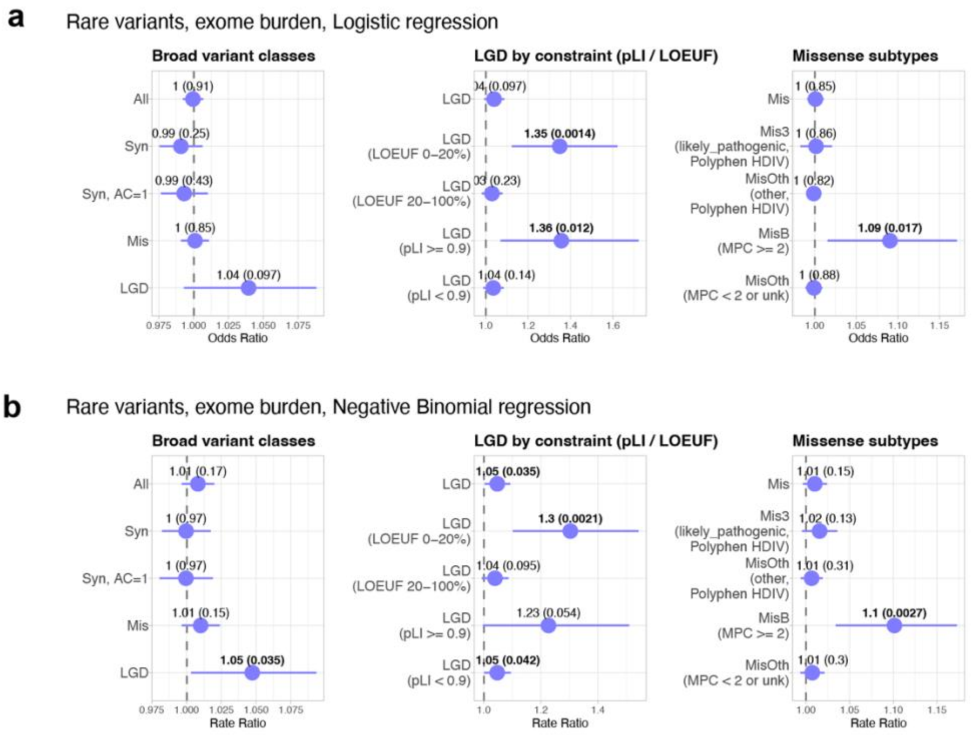


**Supplementary Figure 7 | Rare variant enrichment in European OCD versus control singletons, stratified by genic constraint and missense deleteriousness. a**, Logistic regression testing association between rare variant and case status. **b**, Negative binomial regression models comparing rare variant rates in cases versus controls. Left: broad variant classes (synonymous, missense, and likely gene-disruptive [LGD]). Middle: LGD stratified by genic constraint (pLI, LOEUF; lower LOEUF/higher pLI = greater constraint). Right: missense variants stratified by predicted deleteriousness (Mis3 from PolyPhen-2 HDIV, and MisB from MPC). Odds ratios (ORs; logistic regression) and rate ratios (RRs; negative binomial regression) are shown with 95% confidence intervals. *p* values are two-sided and uncorrected for multiple testing.

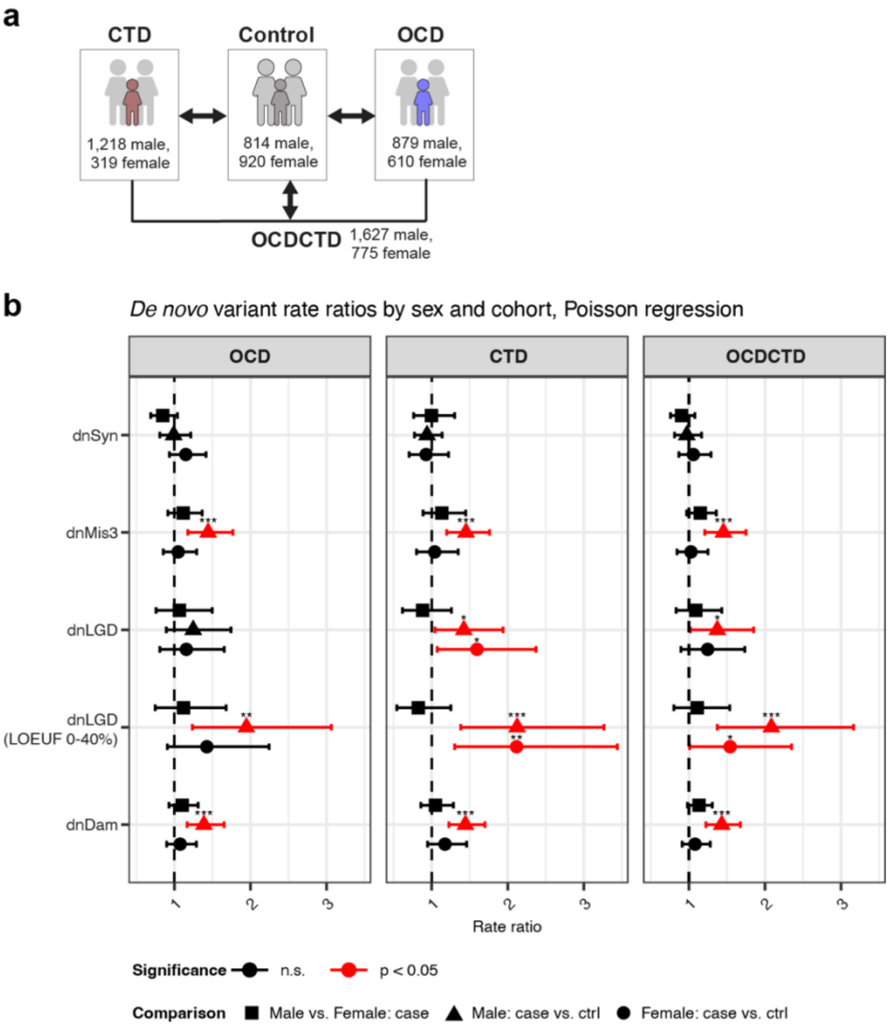


**Supplementary Figure 8 | Sex-stratified *de novo* variant burden across cohorts. a,** Trio proband counts by sex**. b,** Rate ratios (RR) and 95% confidence intervals (CI) for dn variant burden in OCD, CTD, and OCDCTD cohorts, stratified by sex. Poisson regression comparing rates between female cases and female controls (circle), male cases and male controls (triangle), and between male and female cases within each cohort (square). Nominally significant comparisons (two-sided $p < 0.05$) are shown in red. *$p < 0.05$, **$p < 0.01$, ***$p < 0.001$. *P* values are unadjusted for multiple testing.

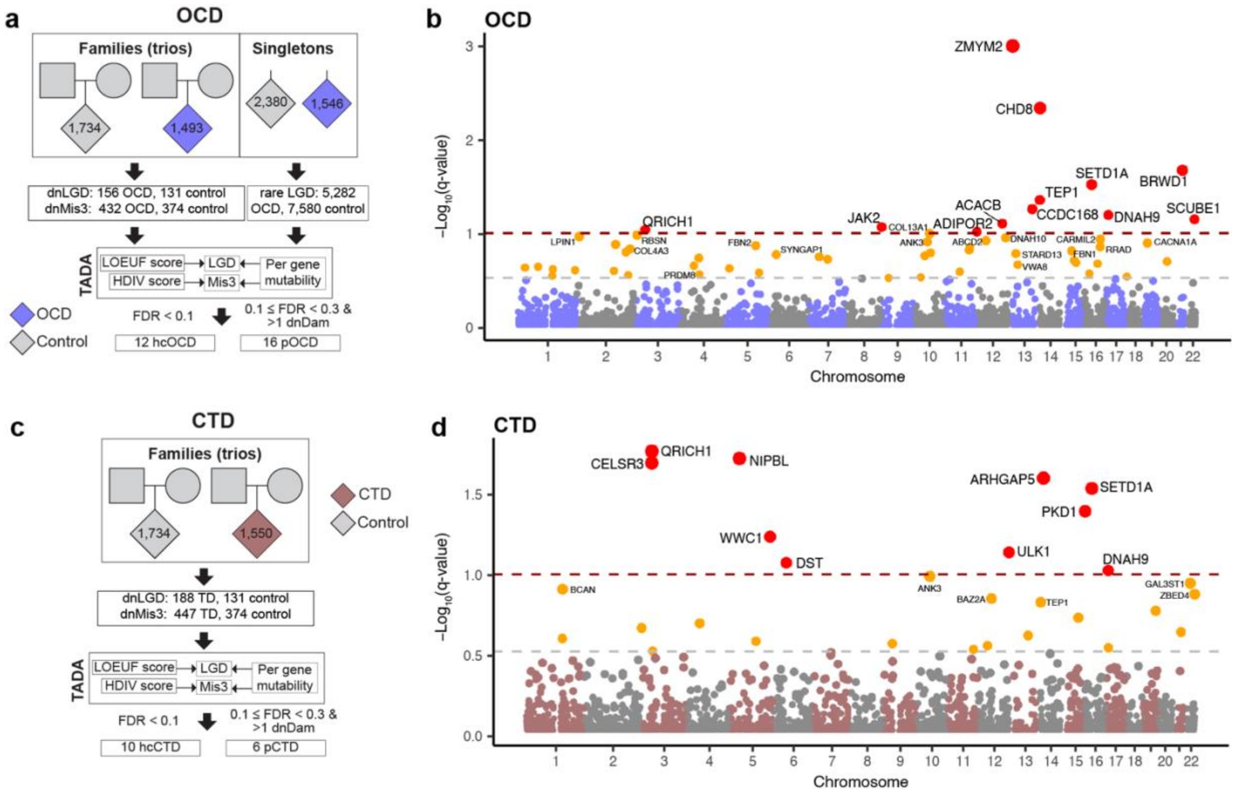


**Supplementary Figure 9 | OCD and CTD gene discovery overview. a,c,** Whole-exome sequencing (WES) data from OCD (a) and CTD (c) were analyzed using a Bayesian framework (TADA), incorporating predicted deleteriousness for likely gene-disrupting (LGD) and missense variants, and gene-specific mutability. Total variants observed per cohort are shown. High-confidence (hc) genes were defined as FDR < 0.1; probable (p) genes as 0.1 ≤ FDR < 0.3 with ≥ 2 dnDam variants. **b,d,** Manhattan plots for OCD (**b**) and CTD (**d**) show log-transformed *q* by chromosomal position. Lines denote FDR = 0.1 and 0.3 thresholds; point color indicates FDR (red: < 0.1, orange: ≥ 0.1 and < 0.3), with labels shown for hc and p genes only.

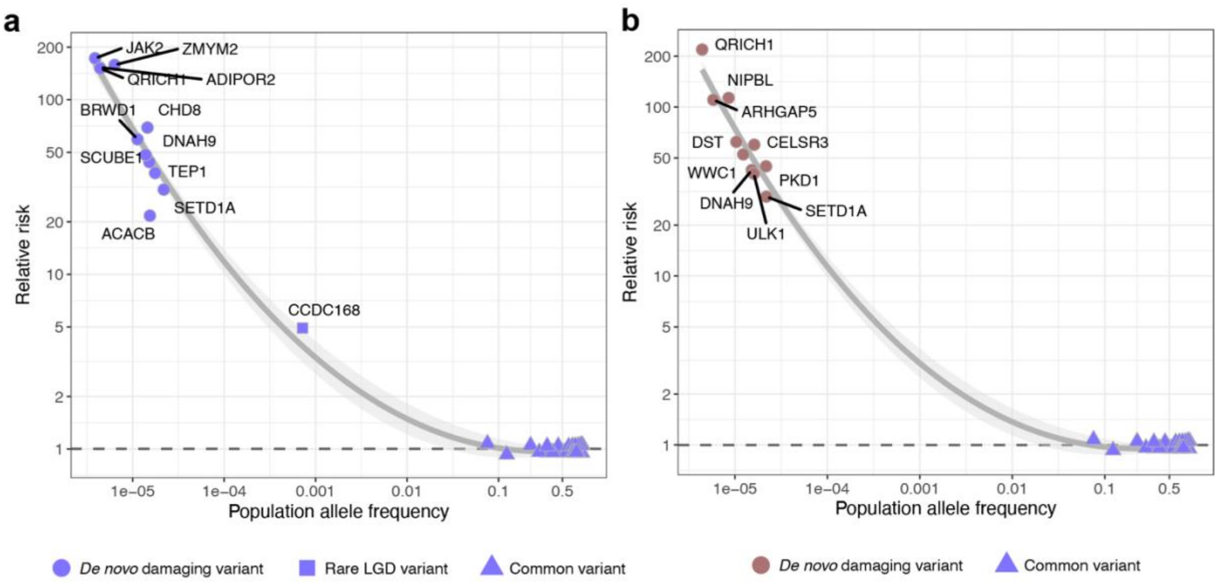


**Supplementary Figure 10 | Gene-level risk architecture for OCD and CTD.** The relationship between population allele frequency and relative risk (RR) is shown for genes and loci implicated in OCD (**a**) and CTD (**b**). Data include high-confidence risk genes identified in this study and OCD GWAS loci[13]. Point shape indicates the variant class used to calculate RR: dnDam (dnLGD + dnMis3), rare loss-of-function (LGD), or common variant. RR was estimated as the case-to-control frequency ratio: the observed/expected rate for dn variants, the observed rate in OCD singletons vs. gnomAD controls for rare variants, and reported GWAS odds ratio for common variants. Each gene is represented by a single RR value, prioritizing evidence in the order: dn > rare > common. A LOESS curve with a 95% confidence band shows the trend between allele frequency and RR.

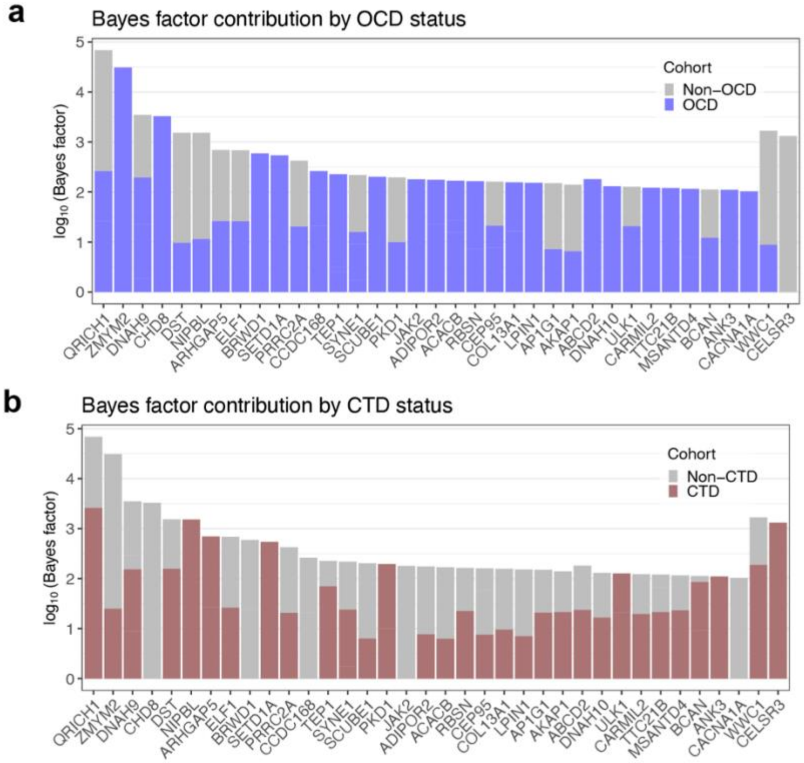


**Supplementary Figure 11 | Gene-level evidence for OCD and CTD association.** Evidence of OCD (**a**) and CTD (**b)** association for each of the 36 high-confidence risk genes.

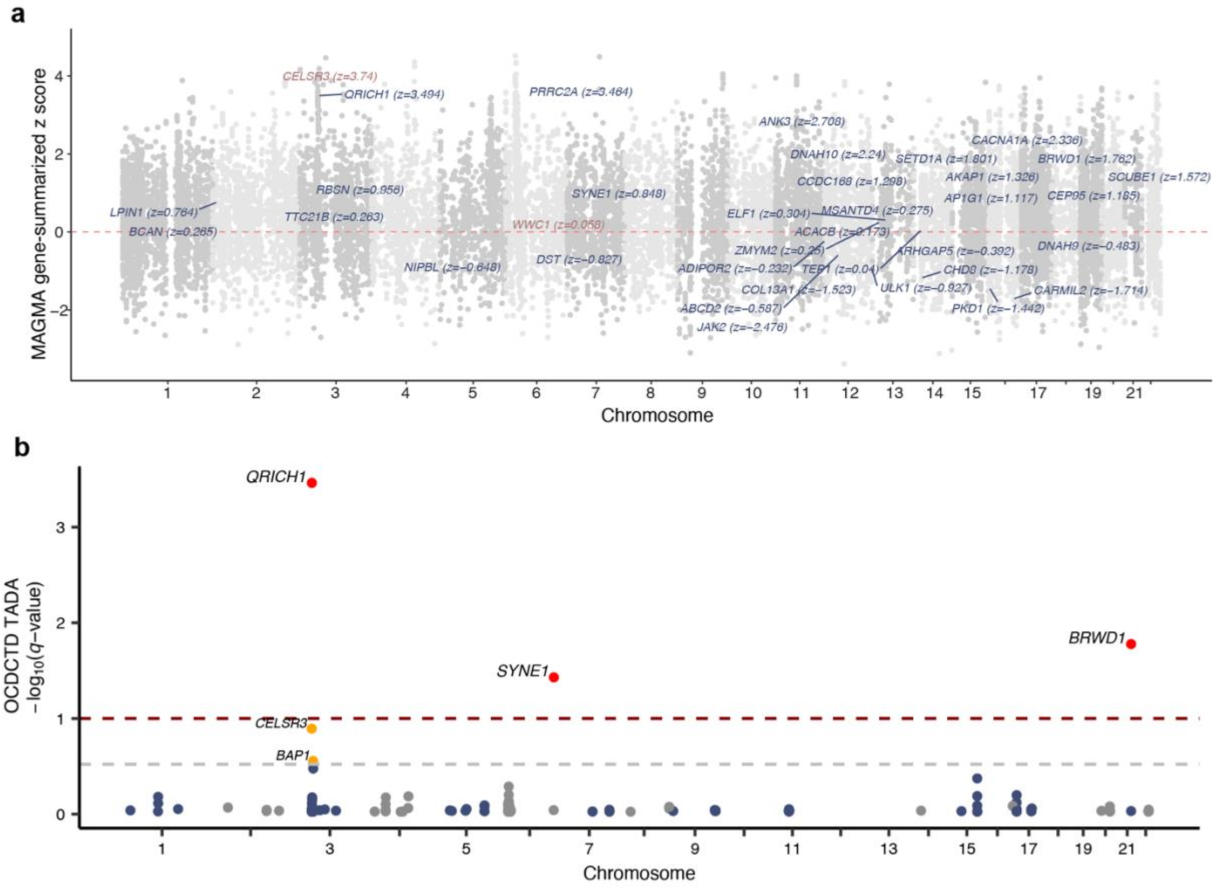


**Supplementary Figure 12 | Overlap of OCD and CTD risk genes with OCD GWAS signals. a,** Gene-level association scores (*z*-scores) were calculated from OCD GWAS summary statistics[13] using MAGMA and plotted by genomic position. hcOCDCTD genes are labelled in dark blue, and two additional hcCTD genes in brown. No genes reach genome-wide significance ($p < 2.5 \times 10^{-6}$; $|z| > 4.75$). **b,** Manhattan plot of TADA *q*-values for OCDCTD, restricted to the 266 genes that were functionally or positionally mapped to significant OCD GWAS loci[13].

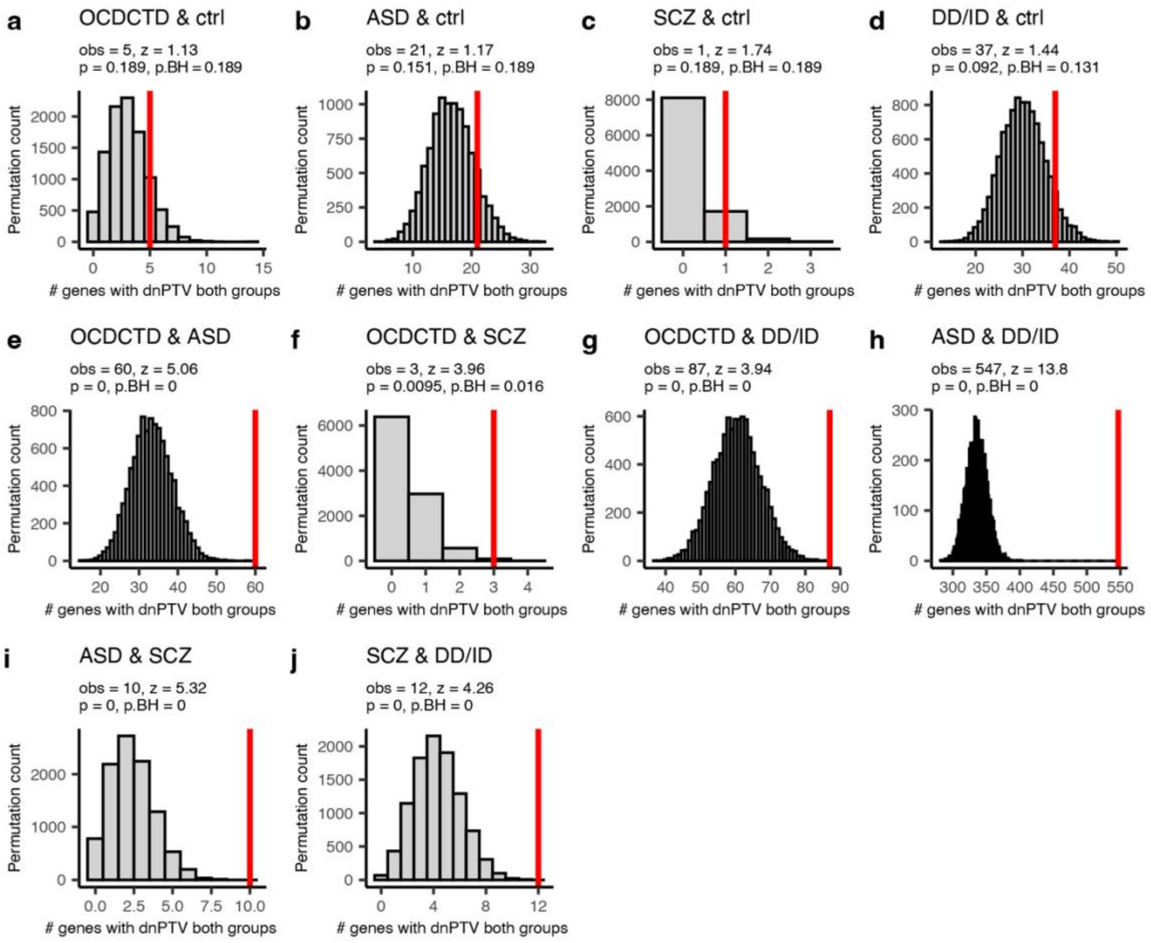


**Supplementary Figure 13 | Cross-disorder dnLGD overlap.** Permutation-based analysis of overlap in genes carrying dnLGD variants across individuals with OCD, CTD, schizophrenia (SCZ), autism spectrum disorder (ASD), and developmental delay/intellectual disability (DD/ID), with unaffected siblings from the Simons Simplex Collection as a negative control (ctrl). **a–j,** Histograms show the null distribution of overlapping genes from 10,000 random permutations (gray); the observed overlap is indicated by a red line. *Z*-scores, empirical *p* values, and Benjamini-Hochberg (BH)-adjusted one-sided *p* values are reported for each pairwise comparison.

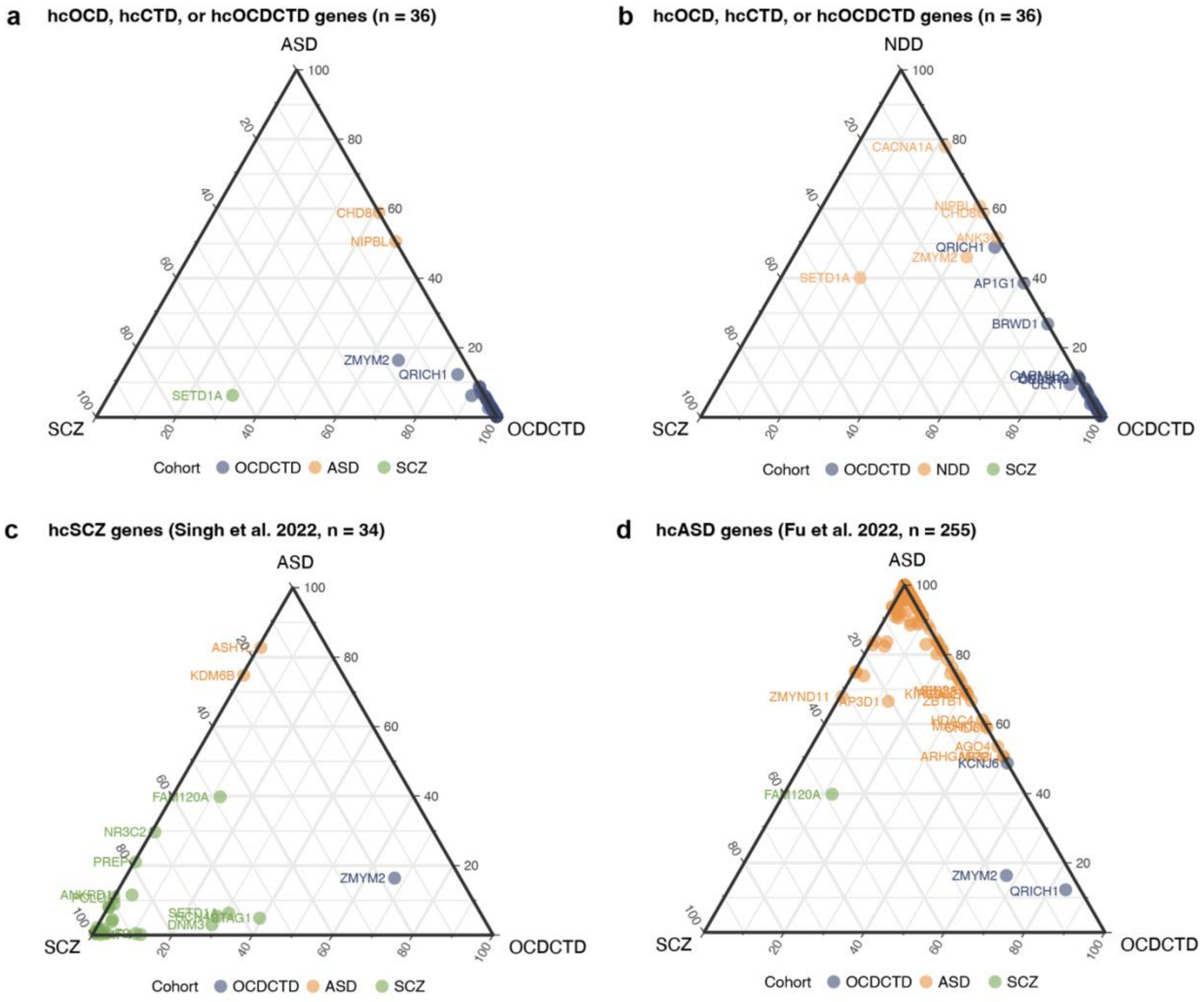


**Supplementary Figure 14 | Relative genetic evidence across neurodevelopmental disorders.** Ternary plots showing relative genetic association strength of risk genes from large whole-exome sequencing studies, based on scaled and normalized $-\log_{10}$($q$-values). Each point represents a gene, positioned according to its relative evidence across the three indicated disorders. **a,b**, n=36 hcOCD, hcCTD, or hcOCDCTD risk genes evaluated across OCDCTD, ASD, and SCZ (**a**), or OCDCTD, NDD, and SCZ (**b**). **c**, n = 34 hcSCZ risk genes[34] evaluated across OCDCTD, ASD, and SCZ. **d**, n = 255 hcASD risk genes[30] evaluated across OCDCTD, SCZ, and ASD. Gene color reflects the disorder with the strongest relative association.

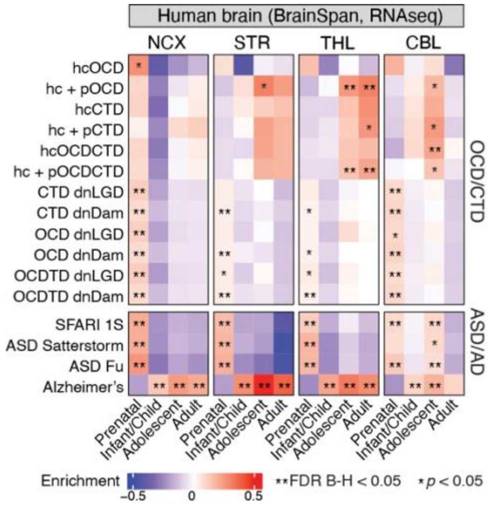


**Supplementary Figure 15 | Enrichment of OCD and CTD risk gene expression in the developing human brain.** Developmental expression of risk gene sets was assessed using BrainSpan RNA-seq data[47], which has a smaller sample size than the BrainSpan microarray dataset[48]. Analyses were performed in four regions: neocortex (NCX), striatum (STR), thalamus (THL), and cerebellum (CBL). Gene expression values were scaled within each region, and heatmaps show enrichment of each gene set relative to all other genes based on one-sided Wilcoxon rank-sum tests. ** Benjamini–Hochberg (B-H) FDR < 0.05; * nominal $p < 0.05$.

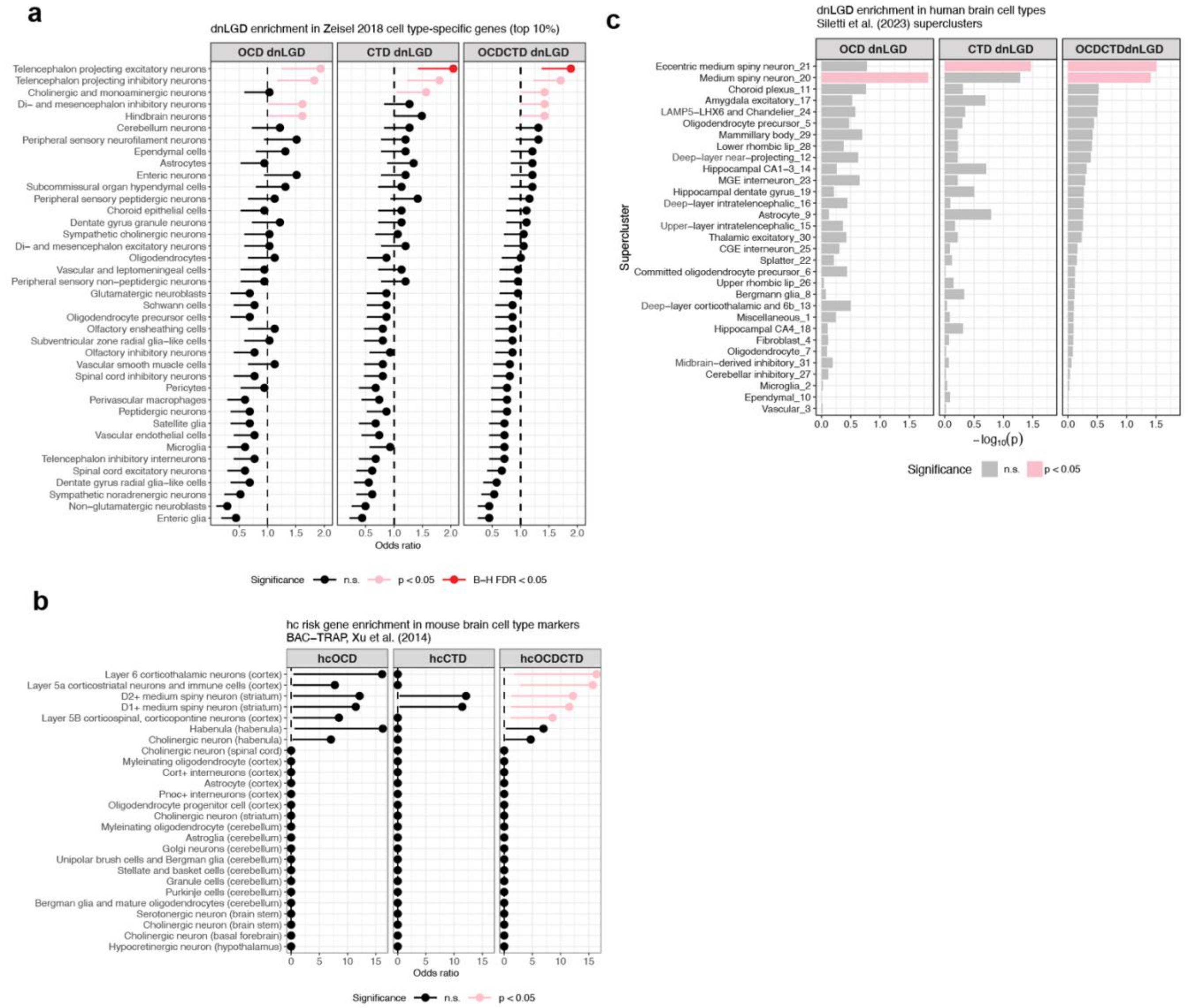


**Supplementary Figure 16 | Enrichment of OCD and CTD risk genes in brain cell types. a,** Enrichment of OCD, CTD, and OCDCTD dnLGD genes in the top 10% cell type–specific genes from 39 broad mouse central and peripheral nervous system cell types[50,55]. Odds ratios and 95% confidence intervals are from one-sided Fisher's exact tests. Pink indicates nominal $p < 0.05$; black indicates not significant (n.s.). No enrichments remained significant after Benjamini–Hochberg (B-H) adjustment. **b,** Enrichment of OCD and CTD risk genes among mouse brain cell type–specific markers defined by BAC–TRAP (bacterial artificial chromosome–translating ribosome affinity purification), which isolates actively translated mRNAs from genetically defined cell populations[51,83]. Odds ratios and 95% confidence intervals were calculated using one-sided Fisher's exact tests. Significance is indicated by color: red (B-H FDR < 0.05), pink (nominal $p < 0.05$), and black (not significant). **c,** Gene set enrichment of OCD/CTD/OCDCTD dnLGD genes using gene rankings by expression specificity in 31 human brain cell types[52]. Bars show $-\log_{10}(p)$ values; pink indicates nominal $p < 0.05$, gray not significant.

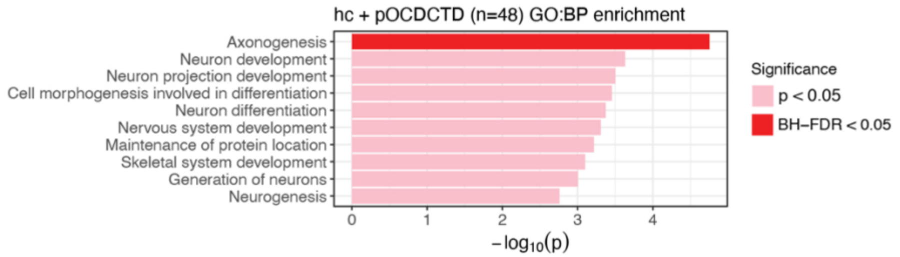


**Supplementary Figure 17 | Gene Ontology enrichment analysis of hc and pOCDCTD genes.** Top 10 most significantly enriched GO:Biological Process terms from overrepresentation analysis of the union of hc and pOCDCTD genes. Pink indicates nominal $p < 0.05$; red indicates significance after Benjamini-Hochberg adjustment (BH-FDR < 0.05).

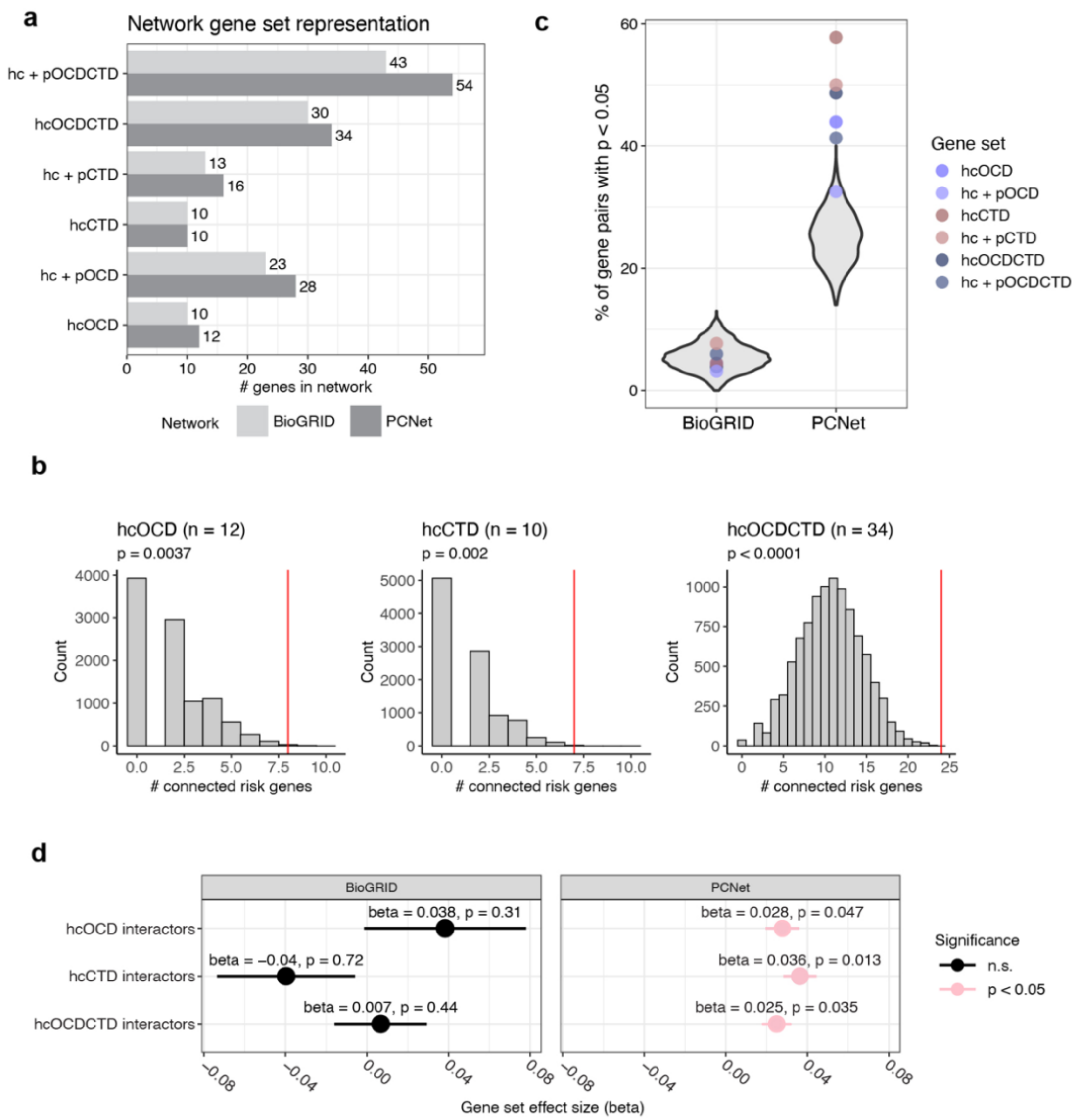


**Supplementary Figure 18 | Risk gene functional connectivity.** Analysis of connectivity among OCD, CTD, and OCDCTD risk genes in two interaction networks. **a,** Gene set representation in BioGRID (physical interactions) and PCNet (multimodal functional interactions). **b,** Histogram of within-set connected genes in 10,000 random gene sets compared to observed risk gene sets in PCNet. Red lines indicate observed values; permutation *p*-values are unadjusted. **c,** Violin plots show the null distribution of the percentage of gene pairs with significant interactor overlap; colored dots indicate observed values for each gene set. **d**, Association of first-degree interactors with OCD common variant risk[13] using MAGMA gene set analysis. Beta (β) indicates effect size; error bars show standard error. Raw *p* values are shown; pink indicates nominal significance ($p < 0.05$).

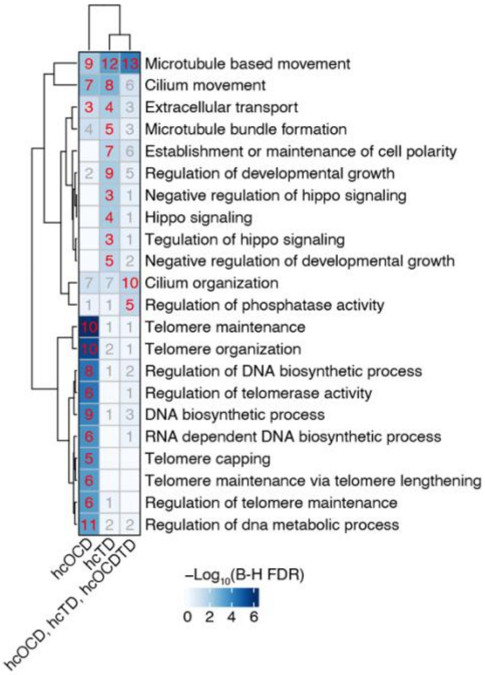


**Supplementary Figure 19 | Functions associated with hc risk genes.** GO:BP overrepresentation analysis of the top 100 genes most connected to hc gene sets via network propagation (hcOCD, hcCTD, and their union with hcOCDTD). Terms were clustered by Jaccard distance. Heatmap color indicates enrichment significance. Text labels indicate the number of hc genes that overlap each term (red: Benjamini-Hochberg [B-H] FDR < 0.05; gray: not significant).

### Supplementary Table Legends

**Supplementary Table 1 | Sample counts by source.**
**Supplementary Table 2 | Sample metadata for all post-QC trios.** 1,734 control trio families and 2,418 OCD or CTD trios, of which 16 trios are published-only (i.e., did not pass sample QC in this study).
**Supplementary Table 3 | Sample metadata for all post-QC singleton samples.** Includes 3,299 control (2,380 European [EUR]) and 2,002 OCD (1,546 EUR) singletons that passed QC. Only the EUR subset was used for burden and TADA analyses.
**Supplementary Table 4 | Number of trio samples excluded by sample QC criteria.**
**Supplementary Table 5 | Number of singleton samples excluded by sample QC criteria**.
**Supplementary Table 6 | All *de novo* variants that were included in burden or TADA analyses.**
**Supplementary Table 7 | Burden test results for *de novo* variants from trio data.**
**Supplementary Table 8 | Burden test results for rare variants from singleton data.**
**Supplementary Table 9 | Overlap of genes affected by *de novo* damaging variants in OCD and CTD subgroups.**
**Supplementary Table 10 | Rare MisB variants in hcOCD, hcCTD, and hcOCDCTD.**
**Supplementary Table 11 | Trio counts by cohort and sex.**
**Supplementary Table 12 | Sex-stratified *de novo* variant burden.**
**Supplementary Table 13 | Estimated number of risk genes.**
**Supplementary Table 14 | Proportion of variants that confer disease risk.**
**Supplementary Table 15 | Proportion of individuals with *de novo* or rare damaging variants contributing to disease risk.**
**Supplementary Table 16 | TADA input and results.**
**Supplementary Table 17 | Number of high-confidence and probable genes.**
**Supplementary Table 18 | Genetic risk architecture.**
**Supplementary Table 19 | Risk gene Bayes factors.**
**Supplementary Table 20 | hcOCD/CTD gene overlap with other risk gene sets.**
**Supplementary Table 21 | Overlap in genes affected by dnLGD variants across neurodevelopmental disorders.**
**Supplementary Table 22 | Scaled genetic association with OCDCTD, ASD, NDD, and SCZ.**
**Supplementary Table 23 | hcOCDCTD enrichment in GTEx tissue-specific markers.**
**Supplementary Table 24 | Brain spatiotemporal risk gene enrichment.**
**Supplementary Table 25 | dnLGD enrichment in mouse brain cell type markers (Zeisel et al., 2018).**
**Supplementary Table 26 | Risk gene enrichment in mouse brain cell type–specific markers defined by BAC–TRAP (Xu et al., 2014).**
**Supplementary Table 27 | dnLGD enrichment in human brain cell type markers (Siletti et al., 2023).**
**Supplementary Table 28 | hc+pOCDCTD GO:BP enrichment.**
**Supplementary Table 29 | Risk gene within-set connectivity by network.**
**Supplementary Table 30 | Risk gene interactome overlap by network.**
**Supplementary Table 31 | Interactor enrichment for *de novo* LGD.**
**Supplementary Table 32 | Interactor enrichment for OCD GWAS-implicated genes.**
**Supplementary Table 33 | Interactor enrichment for OCD common variant risk.**
**Supplementary Table 34 | GO:BP enrichment of top genes from network propagation of hc risk gene sets.**

### Supplementary Notes

### Supplementary Note 1: Study participants

All adult participants and parents of children provided written informed consent along with written or oral assent of their participating child. The Institutional Review Board (IRB) of each participating site approved the study. Pre- and post-QC sample counts by source are summarized in Supplementary Table 1. Supplementary Tables 2 and 3 provide post-QC sample-level information, including OCD and CTD diagnostic status.

***Previously published CTD samples***

We used 511 CTD trios (affected child and both parents) described in our previous "CTD Phase 1" study[22], and 291 trios described in our previous "CTD Phase 2" study[22,23] which included samples from three independent collaborative groups: the Tourette International Collaborative Genetics group (TIC Genetics), the Tourette Syndrome Association International Consortium for Genetics (TSAICG), the Tourette Syndrome Genetics: Southern and Eastern Europe Initiative (TSGeneSEE), and the Uppsala Tourette Cohort (UTC). Ascertainment of these samples have been described previously and are described briefly below:

*TIC Genetics*: Ascertainment of these samples have been described previously[85]. Probands and family members were evaluated for tic disorders, OCD, and ADHD based on the DSM-IV-TR. Probands were required to meet criteria for CTD or another chronic tic disorder. Samples included parent-child trios from simplex and multiplex families.

*TSAICG:* ascertainment of this cohort has been described previously[101]. Inclusion criteria required a TS Classification Study Group (TSCSG) diagnosis of definite TS (a DSM-IV-TR diagnosis of TS plus tics observed by an experienced clinician).

*TSGeneSEE*: Ascertainment of these samples have been described previously[102]. CTD was ascertained according to DSM-III-R, DSM-IV-TR, or DSM-IV criteria, with age of onset before 21 (DSM-III-R) or 18 (DSM-IV and DSM-IV-TR),

*UTC:* Ascertainment of these samples have been described previously[23]. The UTC was collected as part of a study in Sweden named, "Mapping of Hereditary Factors in Neuropsychiatric Conditions, Focusing on Tourette Syndrome." Inclusion criteria required individuals to meet the DSM-IV criteria for CTD.

***New CTD samples***

In this study, we sequenced an additional 2,640 samples (925 probands) through TIC Genetics. TIC Genetic families were ascertained as previously described[85]. The majority of probands were either CTDonly or OCDwCTD (n = 473 and 412, respectively).

***Previously published OCD samples***

*Cappi et al. 2020:* Ascertainment of these samples have been described previously[24]. In brief, this dataset consists of n = 222 patient-child trios in which the offspring met criteria for the diagnosis of OCD, as defined by the DSM-IV-TR or DSM-5. Subjects were assessed using the Structured Clinical

Interview for DSM Axis I Disorders. Subjects with diagnoses of schizophrenia, schizoaffective disorder, autistic disorder, pervasive developmental disorder not otherwise specified, or intellectual disability were excluded. Other diagnostic criteria included onset of symptoms before 18 years of age, no previously diagnosed neurological disorder or OCD occurring exclusively in the context of depression, and no known history of OCD in first-degree relatives.

*Halvorsen et al. 2021 (OCD Collaborative Genetic Association Study, OCGAS):* Ascertainment of these samples have been previously described[25]. Individuals with OCD met DSM-IV diagnostic criteria. A subset of individuals was evaluated for tics, skin-picking and trichotillomania (data reported in Halvorsen et al. 2021[25], annotations were acquired directly from Matthew Halvorsen).

***New OCD samples***

We sequenced 211 OCD parent-child trios (633 individuals) and 1,377 OCD singleton cases through the Foundation for OCD Research. This encompassed samples from multiple sites:

*McLean (PI: Kerry Ressler)*: 1,235 singleton probands were identified through the Mass General Brigham Biobank (MGBB), a biorepository that recruits participants from affiliated hospitals, including McLean Hospital, Massachusetts General Hospital, and Brigham and Women's Hospital[103]. Participants provided informed consent to allow access to their de-identified electronic health records (EHR), donated blood samples, and completed health surveys. Clinical data, including diagnoses, were extracted from the MGBB Portal database. OCD diagnosis was based on the presence of relevant ICD-10 codes in the electronic health record (e.g., based on clinician judgment). Individuals with co-occurring Tourette disorder, autism spectrum disorder, eating disorders, hoarding disorder, or body dysmorphic disorder were excluded.

*University of Florida (UFL, PI: Carol Mathews):* Ascertainment of these samples has been described previously[13,104–106]. This dataset consists of 253 samples, including 37 proband-parent trios (111 individuals) and 142 singletons in which the proband met criteria for the diagnosis of OCD, as defined by the DSM-IV-TR or DSM-5. Subjects were assessed using the Structured Clinical Interview for DSM Axis I Disorders, the Kiddie Schedule for Affective Disorders and Schizophrenia, or the MINI International Neuropsychiatric Interview, along with the Yale Brown Obsessive Compulsive Scale. A small number of probands (3 trios and 3 singletons) with probable OCD—those with strong evidence for the diagnosis but lacking full documentation for a definitive diagnosis—were also included; these individuals typically met symptom severity and impairment thresholds. Subjects with diagnoses of schizophrenia, schizoaffective disorder, autism spectrum disorders, or known intellectual disability were excluded, as were those with previously diagnosed neurological disorders other than tic disorders, head trauma with loss of consciousness, or OCD symptoms occurring exclusively in the context of another psychiatric disorder, such as depression or anorexia nervosa. Individuals with concomitant tic disorders, affective or anxiety disorders, or other obsessive-compulsive related disorders were not excluded.

*Yale (Yale University, PI: Tom Fernandez):* Trios with OCD and/or tic disorders were recruited and sequenced at Yale and identified from several different collections. 21 families were recruited as part of an ongoing genetic study focused on recruiting trios where the proband had a diagnosis of trichotillomania and/or excoriation disorder. For this analysis, probands who self-reported a co-occurring diagnosis of OCD and/or tic disorders were included, and those without were excluded. Ascertainment of these samples has been previously described[107,108] and approved by the Institutional Review Board (IRB) at the Yale School of Medicine. Three additional families where the proband had a clinical diagnosis of OCD or Tourette disorder were collected at Yale and approved by the IRB at Yale School of Medicine.

*Columbia (PI: Tom Fernandez)*: Five families where the proband had a diagnosis of OCD were collected at Columbia University and sequenced at Yale.

*Quebec (PI: Tom Fernandez)*: Families where the proband had a diagnosis of OCD were identified through the Genizon Biobank and were sequenced by Genome Quebec (111 samples, 38 probands).

*Brazil (PI: Tom Fernandez)*: Families where the proband had a diagnosis of OCD were recruited at the University of São Paulo School of Medicine and sequenced at Yale (179 samples, 68 probands).

***Previously published control samples***

*Simons Simplex Collection*: We used WES data from 5,455 samples (1,822 families) from the Simons Simplex Collection (SSC)[86–88]. These control trios comprise neurotypical siblings of autism probands from the SSC and their parents. Ascertainment of SSC families has been described previously[35]. Siblings and parents have no evidence of autism spectrum disorder, intellectual disability, or significant language delays, and, in most cases, have no other major psychiatric or neurological disorders. We used SSC neurotypical sibling trios as controls in family-based analyses, and SSC parents of European ancestry as controls in singleton analyses. Of the post-QC families, 14/1,734 (~0.8%) included a trio member diagnosed with Tourette disorder, and 52/1,734 (~3%) included a trio member diagnosed with OCD. These families were retained in analyses, which is conservative and, if anything, may reduce power without introducing false-positive results.

### Supplementary Note 2: Rationale for variant class selection for gene discovery

Prior sequencing studies of NDDs, including OCD and CTD, have shown that rare and dn variants that contribute to risk are damaging to protein function and particularly enriched in genes under strong selective constraint[22–25,29,30,33,34]. We therefore categorized variants by predicted functional impact. To select variants to use for gene discovery, we evaluated three classes of genetic variation: dn variants (trios), transmitted variants (trios), and rare variants (singletons). Within each class, variants were stratified by functional category: synonymous, missense, and likely gene-disrupting (LGD; stop-gained, frameshift, splice donor, or splice acceptor variants). To enrich for deleterious variants, LGD variants were further stratified by gene-level Loss-of-function (LoF) Observed/Expected Upper bound Fraction (LOEUF) scores, where lower scores indicate stronger intolerance to LoF mutations [39], and missense variants were stratified by predicted functional impact[109].

We observed exome-wide increased rates in cases versus controls for damaging dn and rare variants (see below), but not for rare transmitted variants. Accordingly, only dn and rare variants were included in gene discovery.

#### *LGD variants: LOEUF threshold selection and comparison to pLI*

We assessed two widely used gene-level measures of LoF intolerance:

- pLI (Probability of LoF Intolerance)[95], where pLI ≥ 0.9 is considered intolerant of LoF variation.
- LOEUF[39] from gnomAD v4.1 constraint metrics, where lower LOEUF reflects higher intolerance to LoF variation.

Both pLI[25,30,34] and LOEUF[25,30] have been applied in recent WES-based psychiatric gene discovery studies. Unlike the essentially binary pLI[95], LOEUF[39] provides a continuous metric of loss-of-function intolerance, which may increase sensitivity for detecting enrichment in LoF-intolerant genes. Among the 18,111 genes evaluated, 3,140 had pLI ≥ 0.9. Of these, 1,839 fell within the LOEUF 0–10% percentile range and 1,279 within the LOEUF 10–20% range (59% and 41%, respectively).

We ranked all protein-coding genes by LOEUF percentile and evaluated progressively expanding LOEUF bins, starting from the most LoF-intolerant centile (0–1%) up to the full exome (0–100%) in 1% steps. For each cumulative LOEUF bin, we conducted Poisson regression for dn variants and negative binomial regression for rare variants to assess the relative burden of LGD variants in cases versus controls within each LOEUF-defined gene set. In both OCD and CTD cohorts, we observed the strongest dnLGD enrichment in the most LoF-intolerant decile (0–10%). Additional enrichment peaks were observed between the 10–25% and 25–40% ranges (Supplementary Fig. 5a,b). No clear secondary enrichment peaks were observed for rare singleton LGD variants (Supplementary Fig. 5c). Based on these results, we defined the following LOEUF-based thresholds for gene discovery:

- dnLGD variants**:** lowest 0–40% LOEUF (based on empirical enrichment, Supplementary Fig. 5a,b)
- Rare LGD variants**:** lowest 0–20% LOEUF, consistent with the gnomAD recommended cutoff (LOEUF < 0.6, which corresponds 0–20% LOEUF; https://gnomad.broadinstitute.org/news/2024-03-gnomad-v4-0-gene-constraint/) and which includes all genes with pLI ≥ 0.9.

While pLI ≥ 0.9 showed similar magnitudes of enrichment (Supplementary Figs 6–7, Supplementary Tables 7–8), it captured fewer variants. For example, among 403 dnLGD variants across all trios, 251 had LOEUF 0–40% versus 131 with pLI ≥ 0.9; among 12,862 rare LGD variants across all European singletons, 825 had LOEUF 0–20% versus 553 with pLI ≥ 0.9.

***Missense variants: Mis3 vs. MisB***

We evaluated two missense pathogenicity classifiers: PolyPhen-2 HDIV[38] and MPC[96], both of which have been widely used for WES-based gene discovery in neurodevelopmental and neuropsychiatric disorders[22–25,29,30,34]. We focused on their most deleterious tiers: Mis3 (PolyPhen-2 HIDIV score ≥ 0.957, predicted "probably damaging") and MisB (MPC ≥ 2).

Mis3 variants were prioritized for gene discovery analyses based on:

1. Consistent significant enrichment in dn burden analyses across OCD, CTD, and OCDCTD cohorts in both Poisson and logistic models (Supplementary Fig. 6, Supplementary Table 7),
2. Precedence in OCD[24,25] and CTD[22,23] gene discovery studies, and
3. Greater power for discovery (1,004 of 2,345 dn missense variants were Mis3 versus 222 MisB).

In contrast, MisB variants showed inconsistent enrichment in dn analyses but were significantly enriched in singleton rare variant analyses in OCD (Supplementary Fig. 7, Supplementary Table 8). Based on these findings, we focused on Mis3 for primary gene discovery. Notably, 11 of 36 high-confidence risk genes (hcOCD, hcCTD, or hcOCDCTD) harbored at least one rare MisB variant in OCD singletons—more than expected by chance (OR = 2.16, $p = 0.0003$; Fisher's exact test, one-sided). These rare MisB variants are listed in Supplementary Table 10.

### Supplementary Note 3: Interpretation of high-confidence genes with atypical rare LGD patterns and those unique to the CTD cohort

In our gene discovery analysis, six hc genes had higher observed rates of rare LGD variants in controls than in OCD cases. This pattern modestly influenced the overall TADA results: for *ABCD2*, *CELSR3,* and *WWC1,* it slightly reduced evidence for association, whereas for *DNAH9*, *DNAH10***,** and *TEP1*, it slightly increased the association signal (Supplementary Table 16).

For *ABCD2, CELSR3*, and *WWC1*, singleton controls carry 1-2 rare LGD variants while none are observed in cases. In these instances (cases = 0, controls = 1–2), the likelihood under the alternative hypothesis ($H_1$)—which assumes the gene is a risk gene and expects enrichment of damaging variants in cases—is lower than under the null hypothesis ($H_0$), which assumes the gene is not a risk gene and that variant counts in cases and controls follow the background expectation. This yields a Bayes factor (BF) < 1, and modestly reduces evidence for association. As a result, *CELSR3* and *WWC1* were identified as hcCTD genes but did not meet the FDR < 0.1 threshold in the combined OCDCTD analysis (singleton data were included in the OCDCTD model but not in the CTD analysis). The presence of rare damaging variants in singleton controls modestly reduced the relative signal for these genes, leading to slightly higher FDR values in the OCDCTD model (*CELSR3*: FDR = 0.128; *WWC1*: FDR = 0.116). Gene-level BFs displayed in Fig. 3 and Supplementary Fig. 11 do not show negative evidence from control dnLGD variants, resulting in slightly higher visualized BFs for these three affected genes.

In contrast, for *DNAH9*, *DNAH10,* and *TEP1*, larger counts were observed in both cases and controls (cases: 4-6, controls: 9-12). Here, the $H_0$ likelihood is small given the observed case counts, and the alternative model yields a slightly higher marginal likelihood, producing BF > 1 even though rare LGD rates in controls slightly exceed those in cases. The strength of evidence is modest (BFs between 1.28–3.23 for rare LGD, compared to BFs of 69.9–1326.6 from dnDam variants for these 3 genes). In OCDCTD trio-only TADA analyses, *DNAH9* and *DNAH10* are hc and *TEP1* is a p gene (FDR = 0.011, 0.089, and 0.136, respectively), indicating that the modest contribution from rare variants in controls is not the primary driver of their classification as risk genes.

The singleton control rare LGD variants contributing negative evidence are:

- *CELSR3*: A single rare likely gene-disrupting (LGD) variant was observed in a singleton control (chr3:48639673 C>T), mapping to a splice donor site in intron 34/34 of transcript ENST00000164024. Given its location in the terminal intron, this variant may escape nonsense-mediated decay (NMD), potentially attenuating its functional impact.
- *WWC1*: Two rare LGD variants were observed in singleton controls–a stop-gained variant (chr5:168414415 G>T; ENST00000265293), introducing a premature termination codon at position E337* (exon 9/23), and a frameshift variant (chr5:168424023 T>TA; ENST00000265293), resulting in Y589* (exon 11/23).
- *ABCD2*: Identified as a high-confidence gene in the OCDCTD analysis but did not meet significance thresholds in the individual OCD or CTD analyses. A single rare LGD variant was observed in a singleton control (chr12:39553935 T>A; ENST00000308666), introducing a stop-gained mutation at codon K734* (exon 10/10).

As noted above, these control rare LGD variants contributed modest negative evidence, slightly lowering BFs for each gene. Contemporary gene discovery work[30] often implements a floor adjustment to BFs (e.g., setting a minimum BF of 1) so that negative support in certain variant classes does not reduce the overall strength of association; we did not do this here, resulting in a more conservative analysis.

Based on the totality of the evidence, *CELSR3* and *WWC1* are hcCTD and pOCDCTD genes. Future analyses in larger or independent cohorts will help clarify whether they also confer shared risk to both OCD and CTD. Of note, *CESLR3* is one of four genes that overlap with genome-wide significant OCD GWAS loci[13], increasing the likelihood that it contributes to both OCD and CTD phenotypes (Figure 3c). *DNAH9*, *TEP1,* and *DNAH10* were retained as hc genes, supported primarily by dn evidence.

### Supplementary Note 4: Comparison with previously reported risk genes

We replicated previously reported hcOCD genes *CHD8* and *SCUBE1*[24] and hcCTD genes *CELSR3* and *WWC1*[23] based on the same samples and variants. We also identified an additional dnMis3 variant in *WWC1* from an OCDwCTD proband that was not part of our prior CTD-focused studies that identified *WWC1* as an hcCTD[22,23].

Several previously reported risk genes were not supported in our analyses. S*LITRK5*, which approached genome-wide significance in a rare variant study of 1,263 OCD cases[25] and for which knockout in mice leads to OCD-like behaviors[110], was not significant in this study (OCD FDR = 0.95). The prior association was supported by transmitted Mis3 variants from families and rare Mis3 variants from singletons, neither of which were included in our current gene discovery framework because variant rates did not differ significantly between cases and controls (Fig. 1d; Supplementary Fig. 1c). In this study, no dnLGD, dnMis3, or rare LGD variants were observed in *SLITRK5*.

*ASH1L* was previously implicated in a WES study of 100 Tourette syndrome (TS) trios[26], though not in prior TADA-based CTD WES studies[22,23], and showed no association in this study (CTD FDR = 0.86). The original signal was based on a single dn missense variant and a nominally significant burden of inherited damaging variants, with subsequent targeted sequencing in an additional 524 TS individuals of East Asian ancestry[26]. In our cohort, no dnLGD or dnMis3 variants were identified in *ASH1L*. This lack of replication may reflect limited power, stochastic variation, or ancestry differences, though we did not observe ancestry-related differences in dn rates (data not shown).

Finally, *NRXN1* and *CNTN6*, previously associated with TS through rare copy number variants (CNVs)[62], were not implicated in our analysis (CTD FDRs = 0.65 and 0.9, respectively), as we did not conduct a CNV analysis in the present study. We observed a dnMis3 variant in *NRXN1* from an OCDwCTD offspring and a rare LGD variant in an OCD singleton (OCD FDR = 0.56, OCDCTD FDR = 0.5). No dnLGD, dnMis3, or rare LGD variants were observed in *CNTN6.*

### Supplementary Note 5: Gene-level MAGMA analysis of OCD GWAS

The largest OCD GWAS meta-analysis[13] included 53,660 OCD cases. Publicly available summary statistics excluded cases ascertained from a consumer-based setting (23andMe, n = 30,167 cases), representing 56% of the total cases. Gene-level Multi-marker Analysis of GenoMic Annotation (MAGMA)[44] showed nominal evidence for common variant association ($|z| > 1.96$, $p < 0.05$) for seven hc genes, including CELSR3 ($z = 3.74$) and QRICH1 ($z = 3.49$), which were also among the four hc genes implicated through functional and/or positional mapping of significant GWAS loci. None of these genes reached the exome-wide corrected significance threshold ($z = 4.75$, reflecting a Bonferroni-adjusted $p < 0.05$ for 18,111 genes) (Supplementary Fig. 12). Nonetheless, the overlap of two hc genes is notable, and the incomplete overlap and modest significance may partly reflect the exclusion of over half the OCD cases from the available GWAS summary statistics.

### Supplementary Note 6: Comparison of gene-level association across disorders

#### *Sources of genetic evidence*

We aimed to compare the relative genetic association of individual genes for different neurodevelopmental disorders. We focused on disorders where rare variant-based gene discovery has been productive, including autism spectrum disorder (ASD), neurodevelopmental disorders (NDD), and schizophrenia (SCZ). The strength of genetic association was quantified using q-values from recent large-scale WES studies:

- **OCD and CTD:** TADA FDR values from the joint OCD and CTD meta-analysis, which included all family-based and singleton samples from the current study.
- **ASD:** TADA FDR values from Fu et al. (2022)[30], which analyzed 49,049 family-based samples (15,036 cases) and 14,188 case-control samples (5,591 cases). FDR q-values were extracted from Supplementary Table 11 (FDR_TADA_ASD column).
- **NDD:** TADA FDR values from Fu et al. (2022), incorporating 46,094 developmental disorder (DD) cases from a joint analysis of ASD cases and an additional 31,058 DD trios reported in Kaplanis et al. (2020)[30,33]. FDR q-values were extracted from Supplementary Table 11 (FDR_TADA_NDD column).
- **SCZ:** Q-values from Singh et al. (2022), which analyzed 24,248 schizophrenia cases and 97,322 controls[34]. Q-values were extracted from Table S5 – Gene Results (Q_meta column).

#### *Transforming and scaling gene-level evidence*

To allow meaningful comparisons across disorders with different sample sizes and studies with differing variant classes (e.g., dn, transmitted, rare) and variant types (e.g., SNVs, indels, CNVs), we applied a two-step normalization procedure to the q-values.

First, q-values were transformed to reflect evidence strength using:

$$x_i = -log_{10}(max(q_i, 10^{-8}))$$

Where $q_i$ is the raw gene-level $q$-value, and the lower bound of $10^{-8}$ avoids infinite values while capping the maximum signal to control for differences in statistical power and mitigate the influence of extreme significance from highly powered studies

Second, the transformed values were min-max scaled to [0.1] within each disorder-specific cohort:

$$\underline{x_i} = \frac{x_i - min(x)}{max(x) - min(x)}$$

This procedure preserves relative evidence strength within each disorder while allowing direct visual and analytical comparison across disorders.

#### *Ternary plot visualization*

To explore the relative contributions of different disorders to gene-level genetic associations, we used ternary plots, triangular diagrams that map three-part compositional data. Each axis represents one component of a three-part ratio summing to 1, so each gene can be plotted as a point reflecting its relative association strength across three disorders (Supplementary Fig. 14; Supplementary Table 22).

For example, a gene with scaled values of 2.0 (OCDCTD), 4.0 (ASD), and 4.0 (SCZ) normalizes to (0.2, 0.4, 0.4), placing it near the ASD–SCZ edge, indicative of strong signal in both disorders. Conversely, a gene with a high OCDCTD-specific signal (e.g., 4.0 for OCD/CTD and 0.1 for ASD and SCZ) normalizes to approximately (0.95, 0.024, 0.024), placing it at the OCDCTD vertex, reflecting stronger relative association in OCDCTD. Ternary plots were generated using the ggtern R package.

***Interpretation considerations***
These ternary plots summarize the relative strength of association signals observed in current large WES studies rather than biological specificity. Differences in cohort size, study design, and variant ascertainment contribute to the observed patterns. Therefore, ternary plots should be interpreted as a summary of current genetic evidence rather than a definitive map of disorder-specific biology.
